# Effectiveness of 2025-2026 mRNA-1283 and BNT162b2 COVID-19 Vaccines Against COVID-19 Related Hospitalizations and Medically-Attended COVID-19 Among Adults Aged ≥ 65 years in the United States

**DOI:** 10.64898/2026.04.13.26350772

**Authors:** Nevena Vicic, Alina Bogdanov, Heather Hensler, Taylor Ryan, Ni Zeng, Ekkehard Beck, Emily Patry, Machaon Bonafede, Andre B. Araujo, Amanda Wilson

## Abstract

**Background:** The 2025–2026 COVID-19 vaccine season introduced updated formulations targeting the LP.8.1 lineage. This study assessed the absolute vaccine effectiveness (aVE) of mRNA-1283 and BNT162b2 on COVID-19 outcomes in adults aged ≥65 years.

**Methods:** This retrospective study used linked electronic health record and administrative claims data through Jan 31, 2026. Adults ≥65 years who received the mRNA-1283 or BNT162b2 2025–2026 COVID-19 vaccine were matched to unvaccinated individuals. Inverse probability of treatment weighting was applied to each vaccine’s matched cohorts to balance covariates. Each vaccine was evaluated independently against its own unvaccinated comparator group. aVE against COVID-19 related hospitalization and medically-attended COVID-19 was estimated using Cox proportional hazards models; aVE = 100 × (1 − hazard ratio [HR]).

**Results:** We identified 233,072 mRNA-1283 recipients and 422,610 BNT162b2 recipients ≥65 years. The aVE (95% confidence interval) of mRNA-1283 against COVID-19 related hospitalization and medically-attended COVID-19 was 59.3% (39.0%, 72.9%) and 42.0% (35.0%, 48.3%) among adults ≥65 years and 66.9% (45.9 %, 79.8%) and 50.2% (42.1%, 57.2%) in ≥75 years, respectively. The aVE of BNT162b2 against COVID-19 related hospitalization and medically-attended COVID-19 was 48.3% (32.4%, 60.5%) and 41.2% (36.2%, 45.8%) in ≥65 years and 45.9% (26.0%, 60.4%) and 44.0% (37.8%, 49.6%) in ≥75 years, respectively.

**Conclusions:** This is the first real-world evidence showing that mRNA-1283 prevents COVID-19-related hospitalizations and medically attended events in vulnerable older adults at highest risk of severe disease. These findings support mRNA-1283 as an important public health tool for reducing the ongoing burden of COVID-19.

## Introduction

COVID-19 continues to pose a substantial public health burden, with older adults experiencing the highest rates of severe disease, hospitalization, and death [1]. In the United States, adults aged ≥65 years account for the majority of COVID-19–related hospitalizations, reflecting immunosenescence and the high prevalence of underlying medical conditions associated with severe outcomes [1,2]. Despite the widespread availability of vaccines, this population remains vulnerable due to waning immunity, ongoing viral evolution, and suboptimal vaccine uptake in recent seasons [3,4].

The continued evolution of SARS-CoV-2 has necessitated annual updates to COVID-19 vaccine formulations to maintain alignment with circulating variants [5]. Real-world evidence from prior seasons has demonstrated that updated COVID-19 vaccines reduce the risk of COVID-19 related hospitalization and severe outcomes but effectiveness may vary by product and population [6–10]. For the 2025–2026 season, updated formulations targeting the LP.8.1 lineage of the JN.1 subvariant family were authorized in the United States [11]. Unlike prior COVID-19 vaccines, which utilized the full-length spike protein, mRNA-1283 (mNEXSPIKE) encodes only the receptor-binding and N-terminal domains of the spike protein [12], key targets of neutralizing antibodies and T-cell responses. The vaccine was approved by the US Food and Drug Administration in May 2025 for the prevention of COVID-19 in adults aged ≥65 years and in individuals 12 – 64 with ≥1 underlying condition that puts them at increased risk for severe COVID-19.

At the time of this analysis, no published real-world absolute vaccine effectiveness (aVE) data were available for either mRNA-1283 or BNT162b2 in the 2025–2026 season. Given that mRNA-1283 and BNT162b2 represented the majority of COVID-19 vaccines administered to adults aged ≥65 years in the United States during this season, generating timely real-world evidence is critical to inform clinicians, public health decision-makers, and vaccination programs. To address this gap, this interim analysis sought to characterize the real-world aVE of the updated 2025–2026 mRNA-1283 and BNT162b2 COVID-19 vaccines against COVID-19 related hospitalization and medically-attended COVID-19 among adults aged ≥65 years in the United States using a harmonized protocol.

## Methods

### Data Source

This study leveraged electronic health record (EHR) data from the Veradigm Network EHR linked to administrative healthcare claims sourced from Komodo Health spanning 23 August 2024 through 31 January 2026. The EHR data are sourced from ambulatory/outpatient primary care and specialty settings, and the claims data include inpatient, outpatient, and pharmacy sources. This integrated dataset has been previously characterized and used previously in COVID-19 epidemiology and VE research [6,10,13,14].

This study was conducted in accordance with the Declaration of Helsinki and relevant guidelines for observational studies. The research involved de-identified data and did not require direct patient involvement. Therefore, specific ethics approval and informed consent were not needed, as the data were anonymized and complied with patient privacy regulations. Access to the data was provided under agreements with the data provider, which ensured adherence to ethical standards, including the protection of patient confidentiality.

### Participants and Study Design

This was a retrospective matched cohort study using a common protocol, data source, eligibility criteria, analytic framework, and endpoints to estimate product-specific aVE among adults ≥65 years in the US who were vaccinated with either the mRNA-1283 or the BNT162b2 2025-2026 formulation (vaccinated cohorts) between 28 August 2025 and 24 January 2026 (intake period) or had no evidence of vaccination with any 2025-2026 seasonal COVID-19 vaccine (eligible for the unvaccinated cohorts). Patients were followed through 31 January 2026. For each vaccinated cohort (mRNA-1283 and BNT162b2), vaccinated patients were matched 1:1 to unvaccinated individuals based on age, sex, race, ethnicity, geographic region, week of last healthcare activity before start of intake period, and evidence of vaccination with a prior season’s vaccine during the pre-index period.

The index date was defined as the date of vaccination among the vaccinated; among the unvaccinated, the matched vaccinated patient’s index date was assigned as the unvaccinated control patient’s index date. To allow for establishment of vaccine-induced immunity, the cohort entry date (CED) was defined as 7 days following the index date. The pre-index period started 23 August 2024, the date the prior season’s COVID-19 vaccines became available + 1 day.

Individuals were eligible for inclusion in the study if they were ≥65 years at the start of the intake period, had evidence of activity in the database during the intake period, and had continuous medical and pharmacy enrollment (with 45-day allowable gaps) from the start of the pre-index period through CED, regardless of prior vaccination history. Patients were excluded if they had missing or conflicting age, sex, or state of residence, evidence of a COVID-19 diagnosis or COVID-19 treatment 90 days prior to index date through CED, received any COVID-19 vaccine other than the index vaccine within 60 days prior to the index date or between the index date and cohort entry date (7 days post-index), or had <1 day of follow-up. The codes used to identify COVID-19 vaccinations are listed in Supplementary Tables 1 and 2.

The mRNA-1283 and BNT162b2 analyses were conducted independently, each with its own matched unvaccinated comparator cohort. Due to patient selection criteria, individuals included in one vaccinated cohort could not be included in the other vaccinated cohort or either of the unvaccinated cohorts.

Because the mRNA-1283 or the BNT162b2 analyses were conducted independently, an unvaccinated individual could be included in both analyses; however, their index date could differ as it would have been assigned during matching.

Subgroup analyses were conducted among adults 75 years or older on the index date.

### Outcomes

The primary outcome of COVID-19-related hospitalization was defined as a hospitalization claim with a diagnosis code indicating COVID-19 in any position. The secondary outcome of medically-attended COVID-19 was defined as at least one claim or record with a diagnosis code indicating COVID-19 in any position or setting (including hospital admissions as in the primary outcome, as well as emergency department visits, urgent care visits, office visits, telemedicine visits, and laboratory results). The diagnosis codes used to identify COVID-19 are listed Supplementary Table 3. Follow-up time for the assessment of the outcomes started on CED. Patients were followed until the first occurrence of the outcome, or other censoring criteria (health plan disenrollment, vaccination with any COVID-19 vaccine during follow-up, or end of study) (Figure 1).

**Figure 1:**
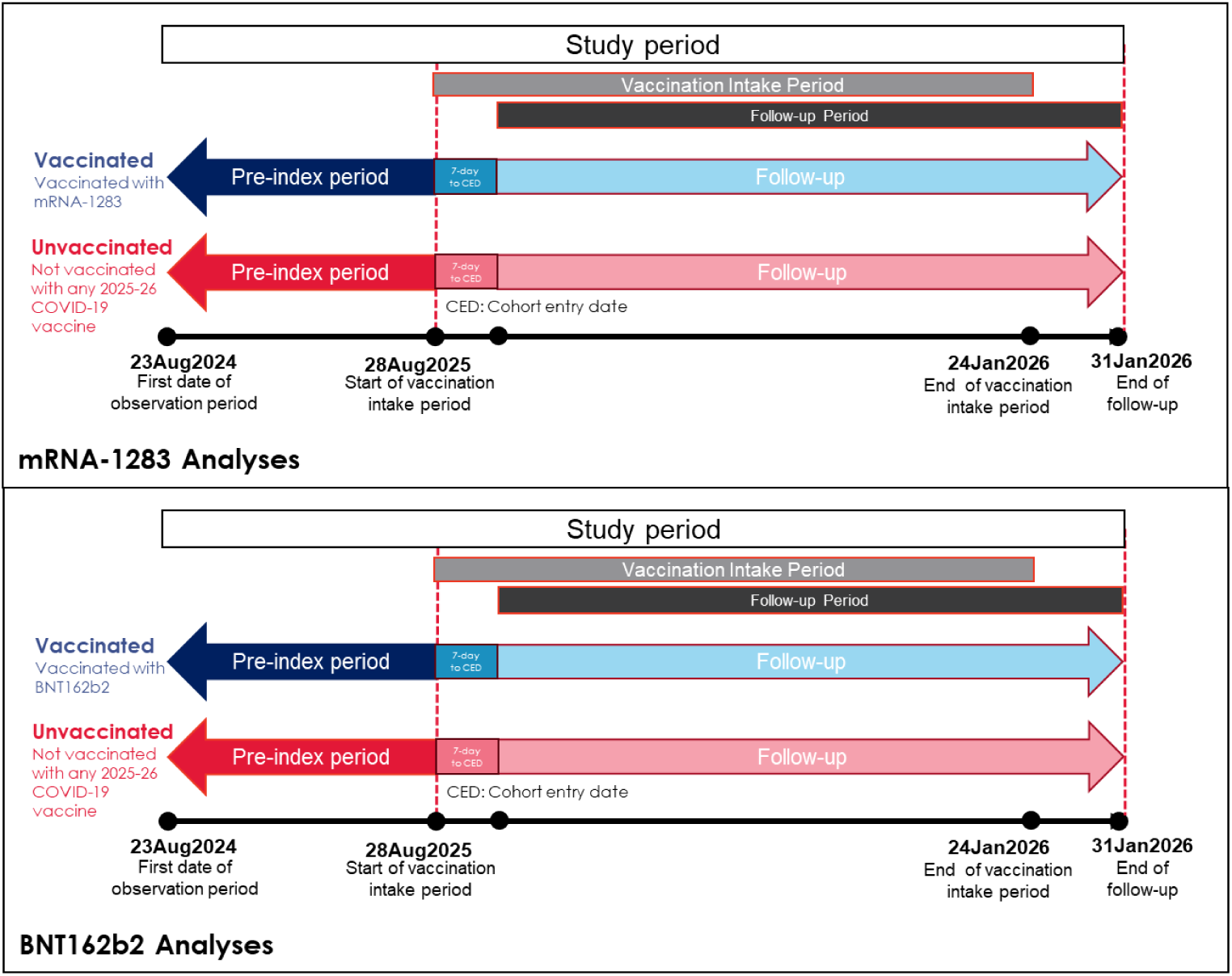
Study Design.

### Covariates

Baseline demographic characteristics captured at the index date included age, sex, race, ethnicity, insurance type, and US geographic region. Clinical characteristics and comorbid conditions [4] associated with high risk for severe COVID-19 outcomes, as defined by the Centers for Disease Control and Prevention (CDC), were ascertained during the 12-month pre-index period and included: asthma, cancer, cerebrovascular disease, chronic kidney disease (CKD), chronic lung diseases, chronic liver diseases, cystic fibrosis, diabetes mellitus (type 1 and type 2), disabilities, heart conditions, HIV, mental health disorders, neurologic conditions, obesity, primary immunodeficiencies, pregnancy, physical inactivity, tobacco use, solid organ or hematopoietic cell transplantation, tuberculosis, use of immunosuppressive medications [2]. Additional baseline variables included: time since last COVID-19 vaccination, time since last COVID-19 infection, week of last healthcare activity prior to the start of the intake period, evidence of prior season (2024–2025) COVID-19 vaccination, evidence of influenza vaccination during the pre-index period, prior diagnosis of influenza during the pre-index period, and healthcare utilization in the per-index period.

Presence of high-risk and immunocompromising conditions were assessed in the pre-index period from up to 365 days prior to the index date, up through the index date (inclusive); pregnancy was assessed from the index date up to 301 days after the index date (inclusive). Healthcare resource utilization was assessed from up to 365 days prior to the index date, up to one day before the index date. Influenza history and influenza vaccination history was assessed from the start of the pre-index period up to one day before the index date. COVID-19 vaccination history was assessed from the start of the pre-index period up to 60 days prior to the index date, and COVID-19 history from the start of the pre-index period up to 90 days prior to the index date.

### Statistical Analysis

The mRNA-1283 and BNT162b2 analyses were conducted independently, including matching procedures and statistical analyses for overall and age-stratified cohorts.

To account for potential differences in measured baseline confounders even after matching, logistic regression models were used to calculate propensity scores and corresponding inverse probability of treatment weights (IPTWs). Stabilized IPTWs were calculated as the probability of being vaccinated to the index vaccine divided by the propensity score (PS) for the vaccinated group and as the probability of being unvaccinated divided by (1-PS) among the unvaccinated group. Covariate balance before and after IPTW was assessed by calculation of the standardized mean differences (SMD). SMDs with absolute values >0.1 indicated covariate imbalance. Covariates were reported descriptively before and after weighting with means and standard deviations for continuous variables and number and percent for categorical variables.

Cumulative incidence and unadjusted HRs were reported for the unweighted sample, and adjusted HRs were reported for the weighted sample. Unadjusted HRs were calculated using a Cox regression model with receipt of index vaccine as the only predictor. Adjusted HRs were calculated for the weighted sample using a weighted Cox regression model including receipt of index vaccine; additional adjustment for covariates with residual imbalance (SMD >0.1) was performed. Cumulative incidence plots with 95% confidence intervals were generated to visually assess the proportional hazards assumption. Subsequently, the unadjusted and adjusted aVE for each outcome were calculated as (1-HR) * 100% and reported with 95% CIs.

Data cleaning and analytic file generation was conducted via SQL. Statistical analyses were conducted using SAS V9.4.

## Results

### Study Population and Baseline Characteristics

The final study population included 233,072 mRNA-1283 recipients and 422,610 BNT162b2 recipients aged ≥65 years who met the patient selection criteria pre-weighting (Figure 2). Baseline characteristics were similar across the mRNA-1283 and BNT162b2 vaccinated cohorts (Supplementary Tables 4 and 5). In the mRNA-1283 analysis, the mean (SD) age of vaccinated patients was 76 (6.4) years, and 56.2% were female. In the BNT162b2 analysis, the mean (SD) age of vaccinated patients was 76 (6.4) years, and 56.3% were female. Most patients in both cohorts had documentation of COVID-19 vaccination during the prior season (mRNA-1283: 84.7%; BNT162b2: 83.2%). Over 90% had their most recent documented activity with the healthcare system within 4 weeks prior to the index date (mRNA-1283: 91.9%; BNT162b2: 92.2%). As these characteristics were all matching criteria, rates were similar in the unvaccinated cohorts that were matched to each vaccinated cohort.

**Figure 2a.**
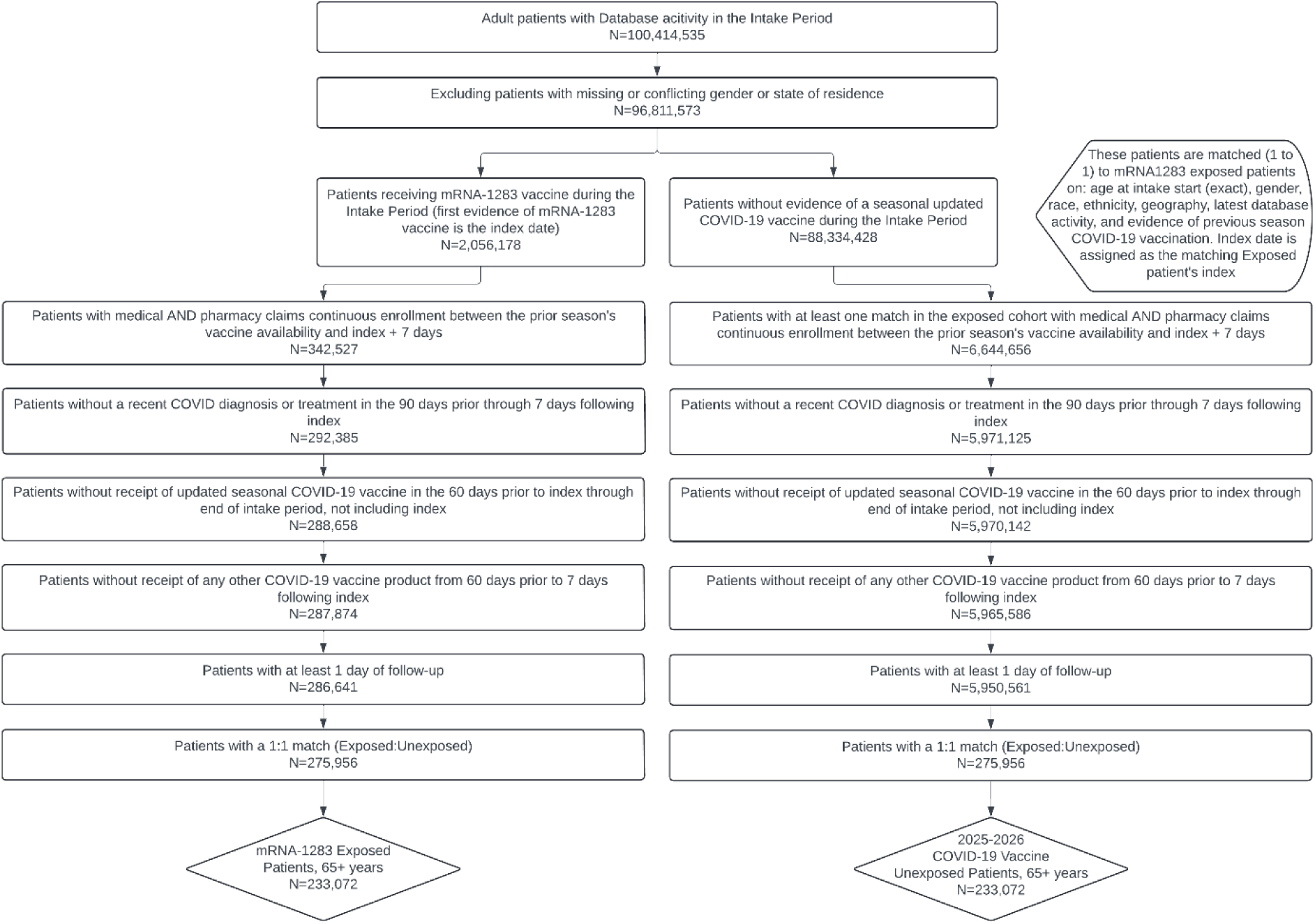
Study Population - mRNA-1283 analysis.

**Figure 2b.**
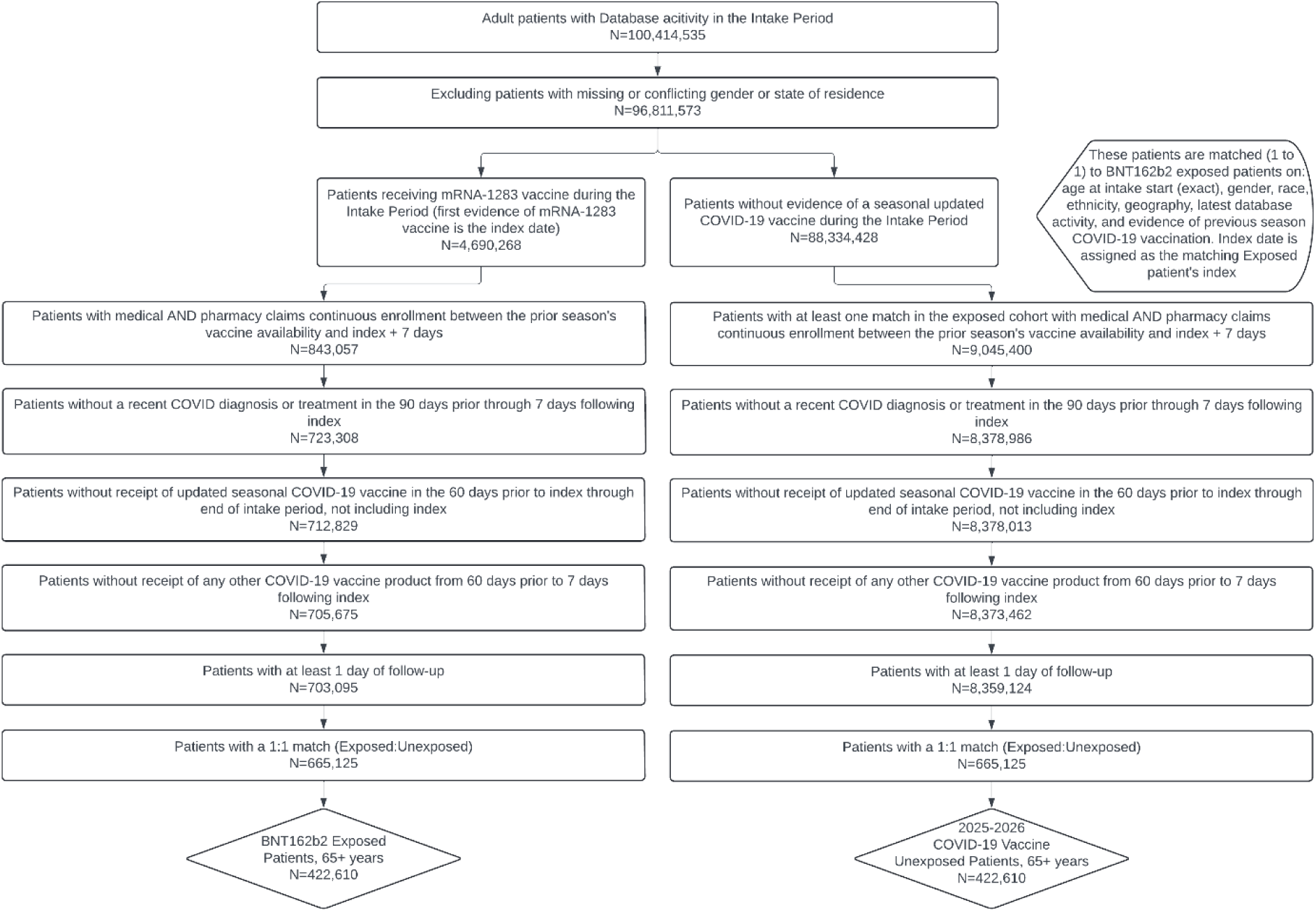
Study Population - BNT162b2 analysis.

Most vaccinations during the study period occurred during October (42%) or September (38%), consistent across both cohorts. The most common underlying medical conditions pre-weighting in the vaccinated cohort were heart conditions (mRNA-1283: 31.7%; BNT162b2: 33.1%), diabetes mellitus (30.7%; 32.9%, respectively), and obesity (26.6%; 28.3%, respectively); corresponding rates in the unvaccinated cohorts were 35.3%, 35.8%, and 29.3% for the mRNA-1283 analysis and 36.0%, 36.5%, and 29.7% for the BNT162b2 analysis. After weighting, baseline characteristics were well balanced between vaccinated and unvaccinated cohorts in both vaccine analyses, with all SMDs ≤0.10.

The median (IQR) duration of follow-up was 69 (42–95) days for mRNA-1283 vaccinated patients and 70 (44–94) days for BNT162b2 vaccinated patients. In the mRNA-1283 vaccinated cohort, 58.6% aged ≥65 and 56.6% aged ≥75 were vaccinated with influenza on the same day as their COVID-19 vaccine; in the BNT162b vaccinated cohort the proportions were 60.3% and 58.2%, respectively.

### Absolute Vaccine Effectiveness Against COVID-19-related Hospitalization and Medically-Attended COVID-19

In the mRNA-1283 analysis, a total of 32 COVID-19-related hospitalizations and 466 cases of medically-attended COVID-19 were identified pre-weighing in the vaccinated, and 83 and 779 in the unvaccinated, respectively (Tables 1 and 2). In the BNT162b2 analysis, a total of 74 COVID-19-related hospitalizations and 931 cases of medically-attended COVID-19 were identified pre-weighing in the vaccinated, and 158 and 1,540 in the unvaccinated, respectively. Cumulative incidence curves for hospitalizations and medically-attended COVID-19 are presented in Figure 3.

**Figure 3.**
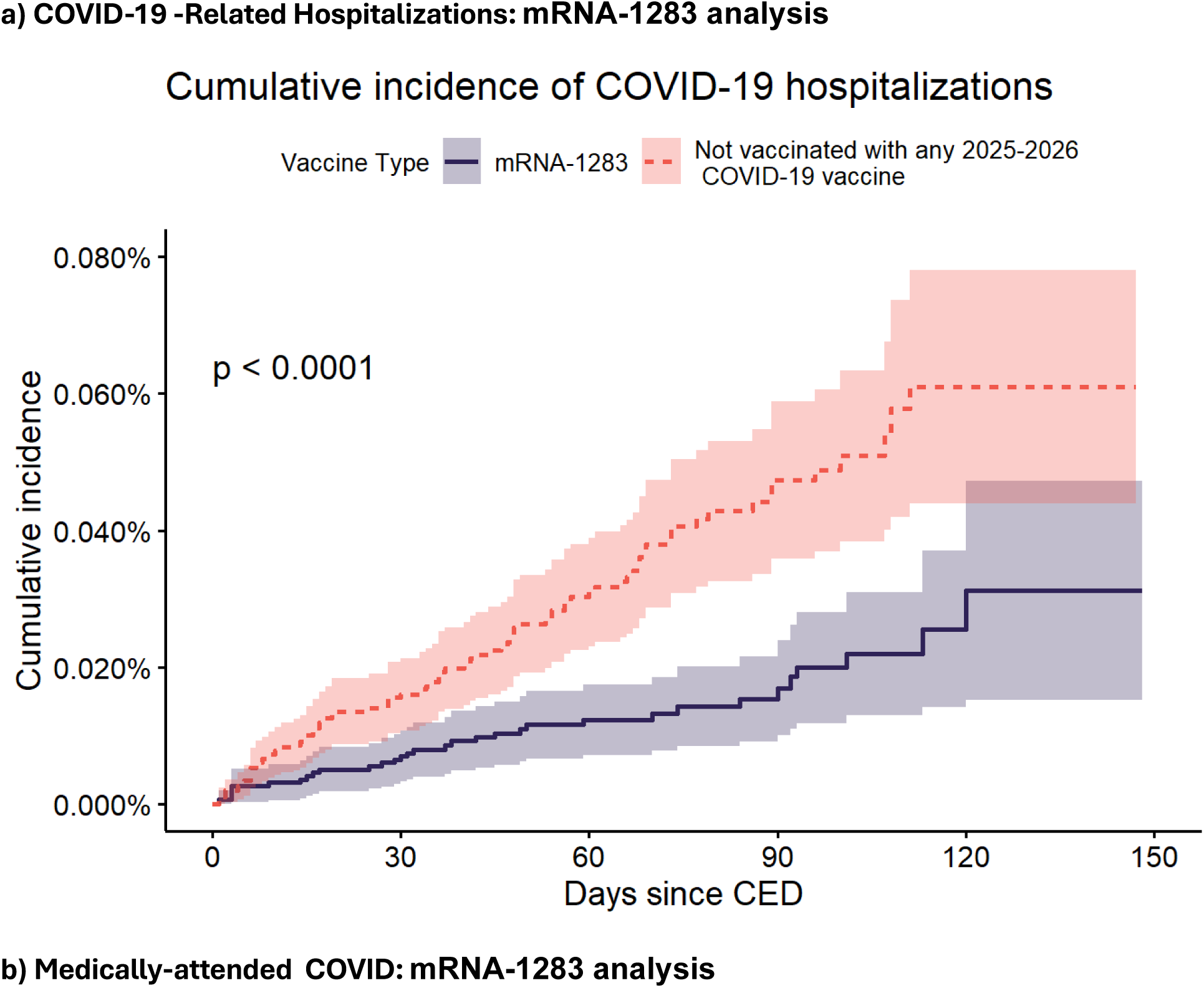
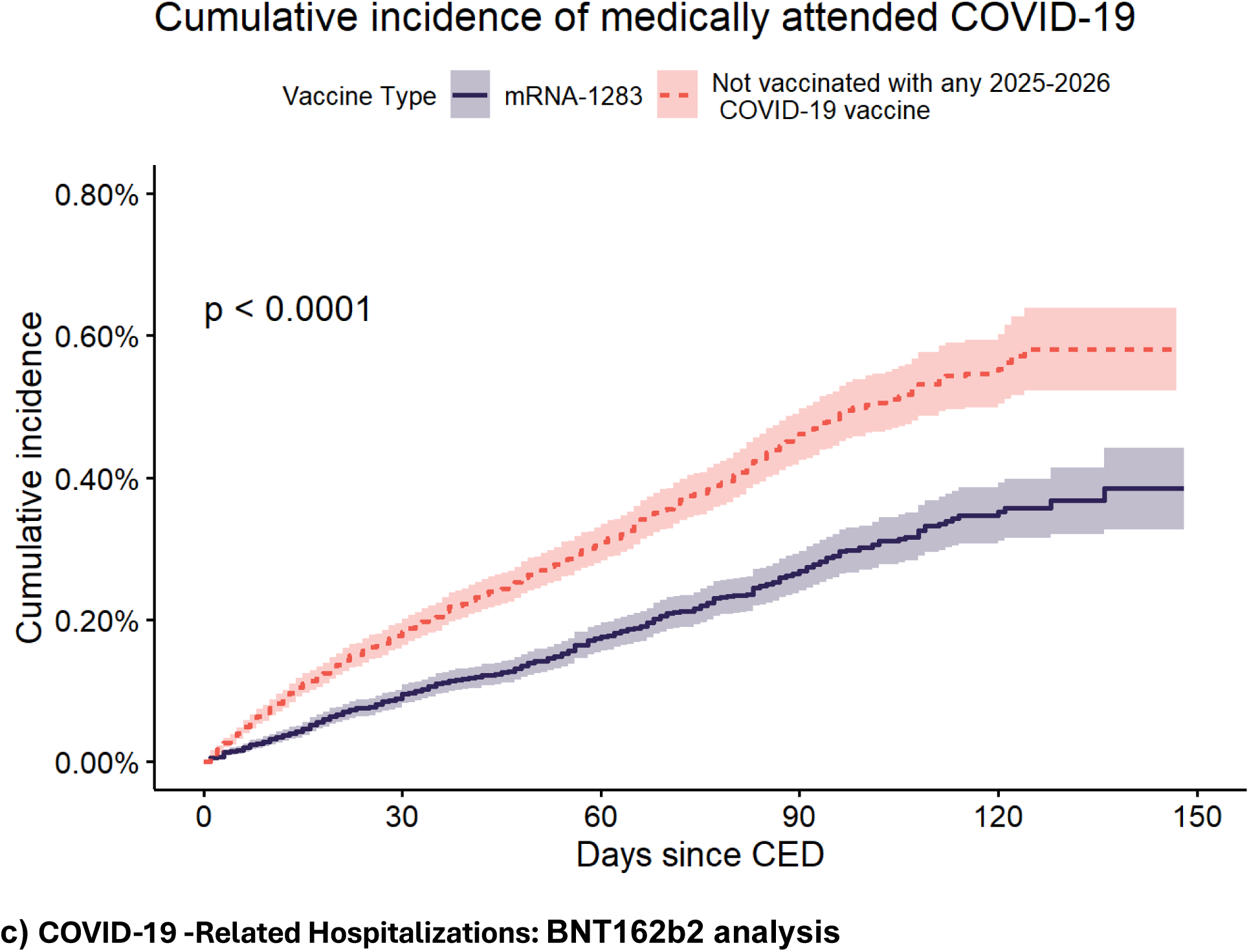
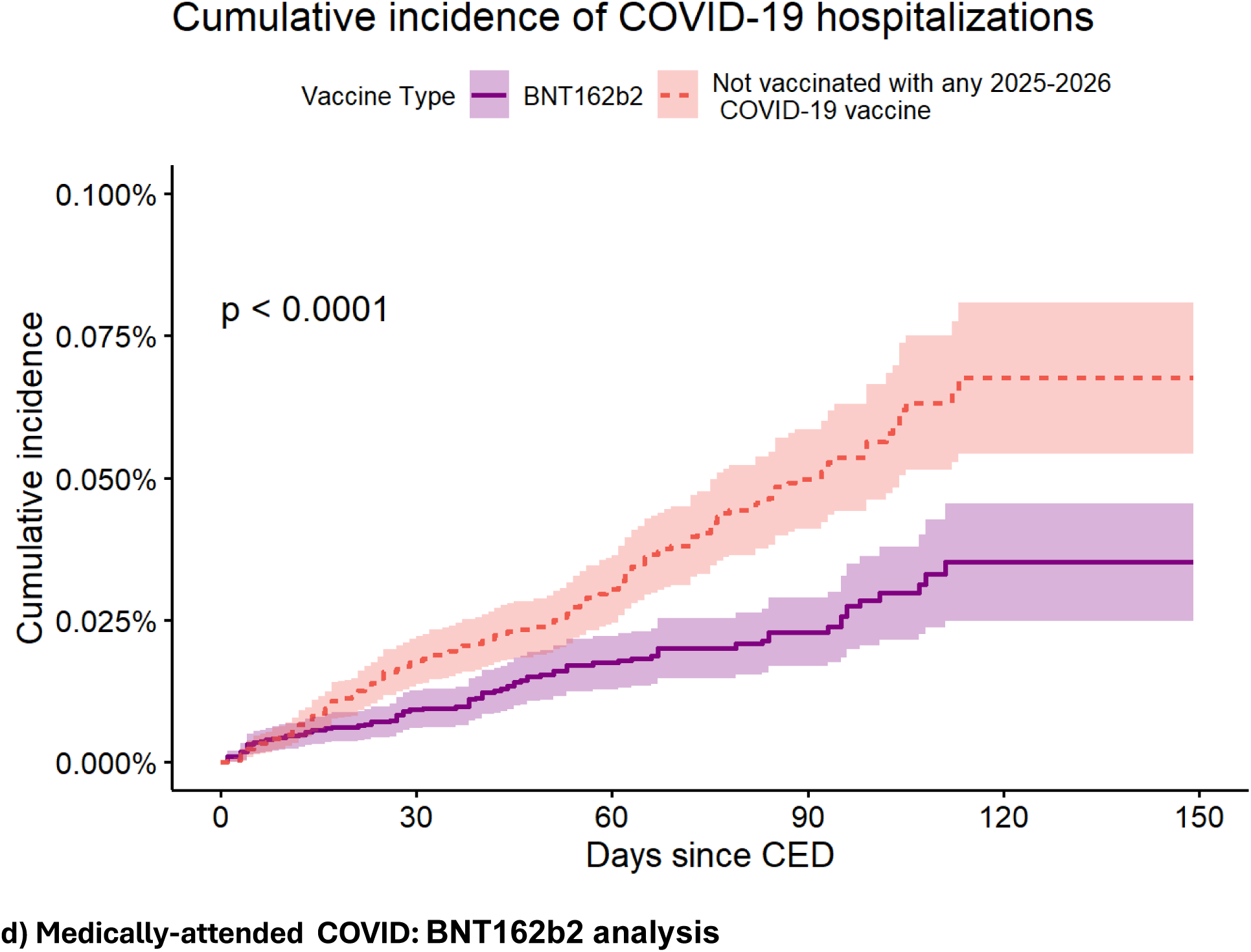
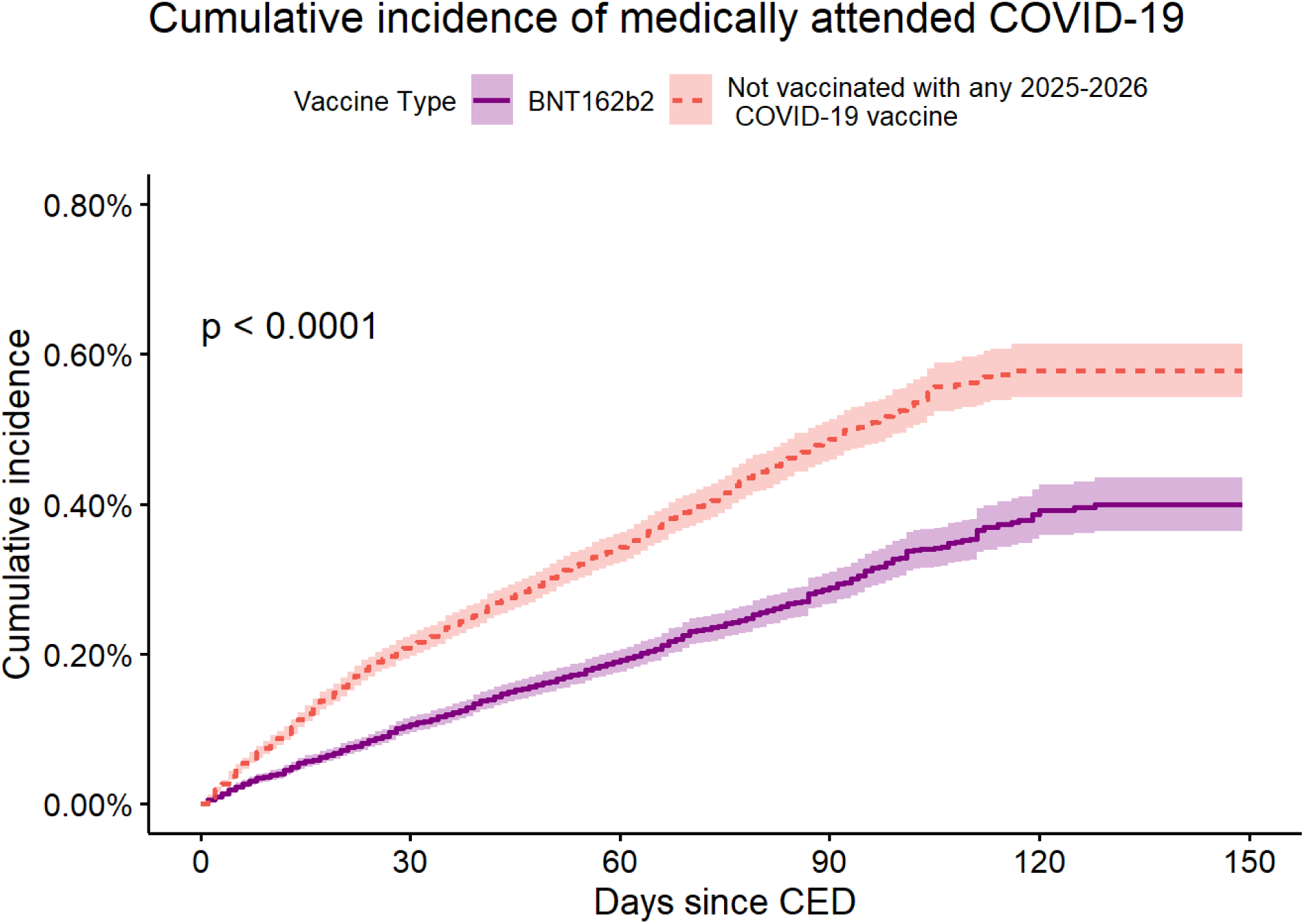
Cumulative incidence curves of time to event.

**Table 1.**
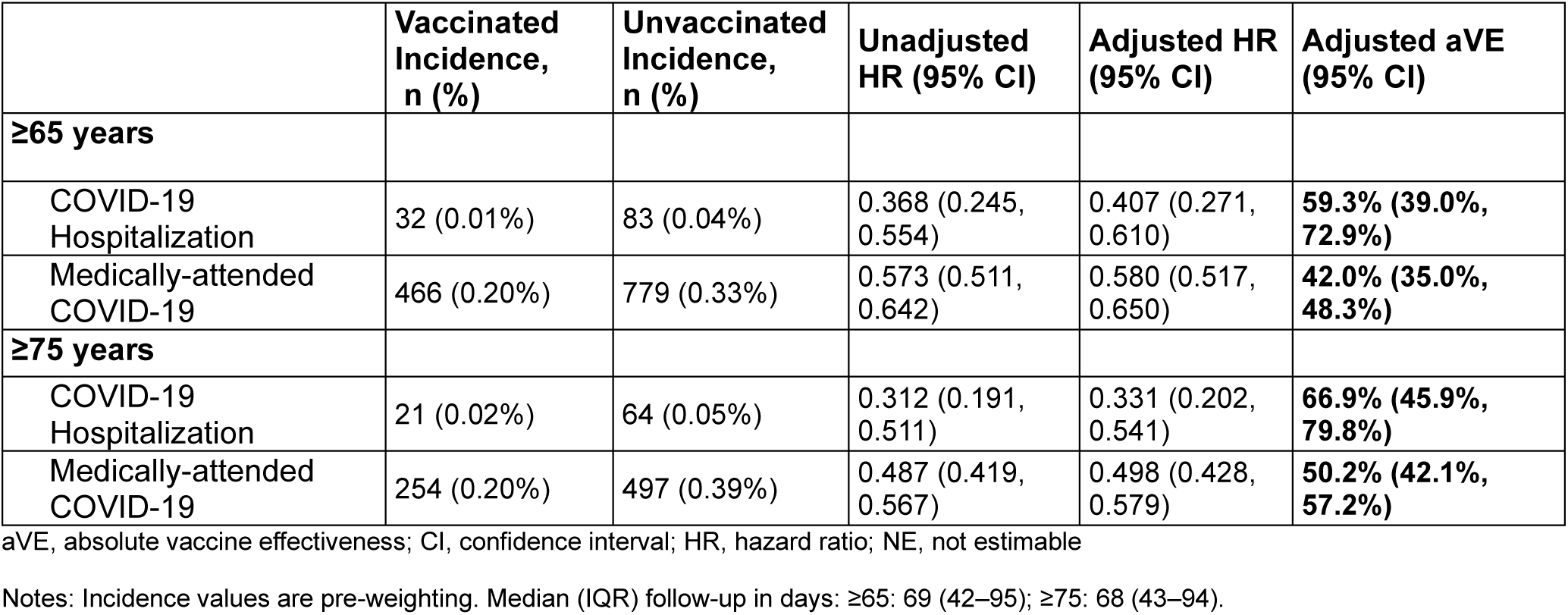
Incidence and Vaccine Effectiveness of COVID-19 -Related Hospitalizations, and Medically-attended COVID-19 for the mRNA-1283 analysis.

|  | Vaccinated Incidence, n (%) | Unvaccinated Incidence, n (%) | Unadjusted HR (95% CI) | Adjusted HR (95% CI) | Adjusted aVE (95% CI) |
| --- | --- | --- | --- | --- | --- |
| **≥65 years** |  |  |  |  |  |
| COVID-19 Hospitalization | 32 (0.01%) | 83 (0.04%) | 0.368 (0.245, 0.554) | 0.407 (0.271, 0.610) | **59.3% (39.0%, 72.9%)** |
| Medically-attended COVID-19 | 466 (0.20%) | 779 (0.33%) | 0.573 (0.511, 0.642) | 0.580 (0.517, 0.650) | **42.0% (35.0%, 48.3%)** |
| **≥75 years** |  |  |  |  |  |
| COVID-19 Hospitalization | 21 (0.02%) | 64 (0.05%) | 0.312 (0.191, 0.511) | 0.331 (0.202, 0.541) | **66.9% (45.9%, 79.8%)** |
| Medically-attended COVID-19 | 254 (0.20%) | 497 (0.39%) | 0.487 (0.419, 0.567) | 0.498 (0.428, 0.579) | **50.2% (42.1%, 57.2%)** |

aVE, absolute vaccine effectiveness; CI, confidence interval; HR, hazard ratio; NE, not estimable
Notes: Incidence values are pre-weighting. Median (IQR) follow-up in days: ≥65: 69 (42–95); ≥75: 68 (43–94).

**Table 2.** Incidence and Vaccine Effectiveness of COVID-19 -Related Hospitalizations, and Medically-attended COVID-19 for the BNT162b2 analysis.

|  | Vaccinated Incidence, n (%) | Unvaccinated Incidence, n (%) | Unadjusted HR (95% CI) | Adjusted HR (95% CI) | Adjusted aVE (95% CI) |
| --- | --- | --- | --- | --- | --- |
| **≥65 years** |  |  |  |  |  |
| COVID-19 Hospitalization | 74 (0.02%) | 158 (0.04%) | 0.441 (0.335, 0.582) | 0.517 (0.395, 0.676) | **48.3% (32.4%, 60.5%)** |
| Medically-attended COVID-19 | 931 (0.22%) | 1,540 (0.36%) | 0.572 (0.528, 0.621) | 0.588 (0.542, 0.638) | **41.2% (36.2%, 45.8%)** |
| **≥75 years** |  |  |  |  |  |
| COVID-19 Hospitalization | 57 (0.03%) | 111 (0.05%) | 0.483 (0.351, 0.665) | 0.541 (0.396, 0.740) | **45.9% (26.0%, 60.4%)** |
| Medically-attended COVID-19 | 546 (0.24%) | 955 (0.42%) | 0.541 (0.487, 0.602) | 0.560 (0.504, 0.622) | **44.0% (37.8%, 49.6%)** |

aVE, absolute vaccine effectiveness; CI, confidence interval; HR, hazard ratio; NE, not estimable
Notes: Incidence values are pre-weighting. Median (IQR) follow-up in days: ≥65: 70 (44–94); ≥75: 70 (45–93).

In the primary analysis of adults aged ≥65 years, both vaccines demonstrated significant aVE against COVID-19 related hospitalization and medically-attended COVID-19 (Tables 1 and 2). After IPTW adjustment, the aVE (95% CI) of mRNA-1283 against COVID-19-related hospitalization was 59.3% (39.0%, 72.9%) and for BNT162b2 it was 48.3% (32.4%, 60.5%). The adjusted aVE (95% CI) of mRNA-1283 against medically-attended COVID-19 was 42.0% (35.0%, 48.3%) and for BNT162b2 it was 41.2% (36.2%, 45.8%).

### Age Subgroup Analyses

In the mRNA-1283 analysis, the vaccinated cohort was comprised of 126,136 adults aged ≥75 years pre-weighting. Baseline characteristics are reported in Supplementary table 6. Among adults aged ≥75 years, aVE against COVID-19 related hospitalization was 66.9% (45.9%, 79.8%) and against medically-attended COVID-19 was 50.2% (42.1%, 57.2%).

In the BNT162b2 analysis, the vaccinated cohort was comprised of 227,989 adults aged ≥75 years pre-weighting. Baseline characteristics are reported in Supplementary table 7. Among adults aged ≥75 years, aVE against COVID-19 related hospitalization was 45.9% (26.0%, 60.4%) and against medically-attended COVID-19 was 44.0% (37.8%, 49.6%).

## Discussion

In this analysis of a large linked EHR and administrative claims dataset, both the 2025–2026 mRNA-1283 and BNT162b2 COVID-19 vaccines demonstrated significant protection against COVID-19 related hospitalization and medically-attended COVID-19 among adults aged ≥65 years in the United States. The aVE against COVID-19-related hospitalization in this population was 59.3% for mRNA-1283 and 48.3% for BNT162b2, while the aVE against medically-attended COVID-19 was 42.0% for mRNA-1283 and 41.2% for BNT162b2. This study provides the first real-world evidence of mRNA-1283 effectiveness.

The targeted design of mRNA-1283, which encodes only the receptor-binding and N-terminal domains of the spike protein, allows for lower mRNA doses compared with COVID-19 vaccines that encode the full-length spike (10 µg for mRNA-1283 vs 50 µg for mRNA-1273 and 30 µg for BNT162b2) [15–17]. mRNA-1283 has been shown to elicit robust immune responses and efficacy in older adults compared with mRNA-1273. These findings are supported by phase 3 clinical data from the NextCOVE trial, in which higher efficacy point estimates were observed for mRNA-1283 compared with mRNA-1273 in individuals at increased risk of severe COVID-19, including older adults (13.5%, 95% CI, −7.7 to 30.6), with larger differences observed in more vulnerable populations, such as older adults with high-risk conditions (28.1%, 95% CI, 4.4 to 45.9) [18]. These data are consistent with improved immune responses, particularly in vulnerable older adult populations.

The public health impact of mRNA-1283 in the United States has also been evaluated in a modeling study, which estimated that compared with no COVID-19 vaccination, mRNA-1283 could prevent approximately 120,000 hospitalizations and 16,360 deaths among adults aged ≥65 years [19].

In this context, because each vaccine cohort was analyzed independently against its own unvaccinated comparator, between-product observations were not conducted in this study and warrant direct comparative evaluation in future studies. With that important limitation, the point estimates for aVE against COVID-19-related hospitalization were numerically higher for mRNA-1283 (59%) than for BNT162b2 (48%) in adults 65 and older with a larger difference observed among adults 75 and older (mRNA-1283: 67%; BNT162b2: 46%). Further evaluation in additional datasets, populations, and with longer follow-up is needed to better characterize durability and to support future comparative effectiveness analyses.

### Limitations

Although we have identified and adjusted for many baseline confounders, cohort assignment was not randomized, and residual confounding may remain, as is inherent to observational study designs. Also, the study population includes individuals engaged with the healthcare system, possibly excluding others due to barriers such as trust, socioeconomic status, or accessibility. The outcome definition of hospitalization with COVID-19 partially mitigates this limitation, as severe outcomes are less influenced by variation in healthcare-seeking behavior. While the study design requires that vaccinated individuals remain event-free for 7 days following vaccination, the use of a symmetric study design among the unvaccinated cohorts minimizes the introduction of bias between cohorts.

As with all claims and/or EHR data, vaccination status may be incompletely captured, which could have led to bias toward the null and result in underestimation of true aVE. Misclassification of both exposure and outcomes is also possible, as diagnostic and procedural codes used to define vaccination and COVID-19–related hospitalizations may be subject to coding errors. However, validation studies suggest that outcome misclassification is likely limited; for example, Kadri et al. reported that the ICD-10-CM code U07.1 for COVID-19 had a positive predictive value of approximately 98% when compared with clinical documentation and laboratory-confirmed infection [20].

Additionally, these data sources may not fully capture all hospitalizations, particularly among populations with limited or fragmented access to care such as uninsured individuals, underrepresented groups, or those receiving care outside the insurance network, which may affect generalizability and introduce selection bias.

### Strengths

This retrospective, observational cohort study implemented a combination of matching and inverse probability weighting to adjust for confounding, specifically the underlying differences between the unvaccinated cohorts and recently vaccinated cohorts. Direct matching on clinical and demographic criteria such as age, sex, race, ethnicity, geographic region, week of last healthcare activity prior to the start of the intake period, and 2024-2025 vaccine receipt was used to assign index dates for the unvaccinated cohorts. Matching on week of last healthcare activity was intended to mitigate potential surveillance bias by ensuring the groups are similar in terms of how often they engage with the healthcare system. The use of IPTW was able to further mitigate confounding and estimate average treatment effect among the full population.

Despite limited sample size, the use of a harmonized protocol, data source, eligibility criteria, analytic framework, and endpoints ensured that individuals in each group, vaccinated and unvaccinated, were handled the same.

## Conclusion

In this analysis of data on the effectiveness of the 2025-2026 mRNA COVID-19 vaccines, both mRNA-1283 and BNT162b2 demonstrated substantial aVE against COVID-19-related hospitalization and medically-attended COVID-19 among adults aged ≥65 years in the United States. Among patients vaccinated with mRNA-1283, numerically higher aVE was observed in patients ≥65 years of age against COVID-19-related hospitalization. These findings provide the first real-world evidence of mRNA-1283 effectiveness in older adults, support continued uptake of updated COVID-19 vaccines, and provide direction for future comparative VE studies.

## Ethical Approval

This study was conducted in accordance with the Declaration of Helsinki and relevant guidelines for observational studies. The research involved de-identified claims data and did not require direct patient involvement. Therefore, specific ethics approval and informed consent were not needed, as the data were anonymized and complied with patient privacy regulations. Access to the data was provided under agreements with the data provider, which ensured adherence to ethical standards, including the protection of patient confidentiality.

## Medical Writing Assistance

Medical writing services were provided by Jessamine Winer-Jones, PhD, an employee of Veradigm.

## Disclosure of AI Use in Manuscript Development

Artificial intelligence–assisted writing tools (Claude, Anthropic) were used to support manuscript drafting, data extraction verification, and document preparation. All content was reviewed, edited, and approved by the authors, who take full responsibility for the accuracy and integrity of the work.

## Authorship

All authors attest they meet the ICMJE criteria for authorship.

## Data Availability

The data that support the findings of this study were used under license from Veradigm and Komodo Health. Due to data use agreements and its proprietary nature, restrictions apply regarding the availability of the data. Further information is available from the corresponding author.

---

**Supplementary Table 1.** List of codes used to identify 2025-2026 COVID-19 vaccines from the Veradigm EHR and linked claims datasets.

| COVID-19 vaccine type | CPT | CVX | NDC |
| --- | --- | --- | --- |
| mRNA-1283 | 91323 | 334 | 80777-0400-17, 80777-0400-60, 80777-0400-61, 80777-0400-62 |
| BNT162b2 | 91319, 91320 | 309, 310 | 00069-2501-01, 00069-2501-10, 00069-2528-01, 00069-2528-10 |

CPT, Current Procedural Terminology; CVX, vaccine administered codes; NDC, National Drug Codes

**Supplementary Table 2.**
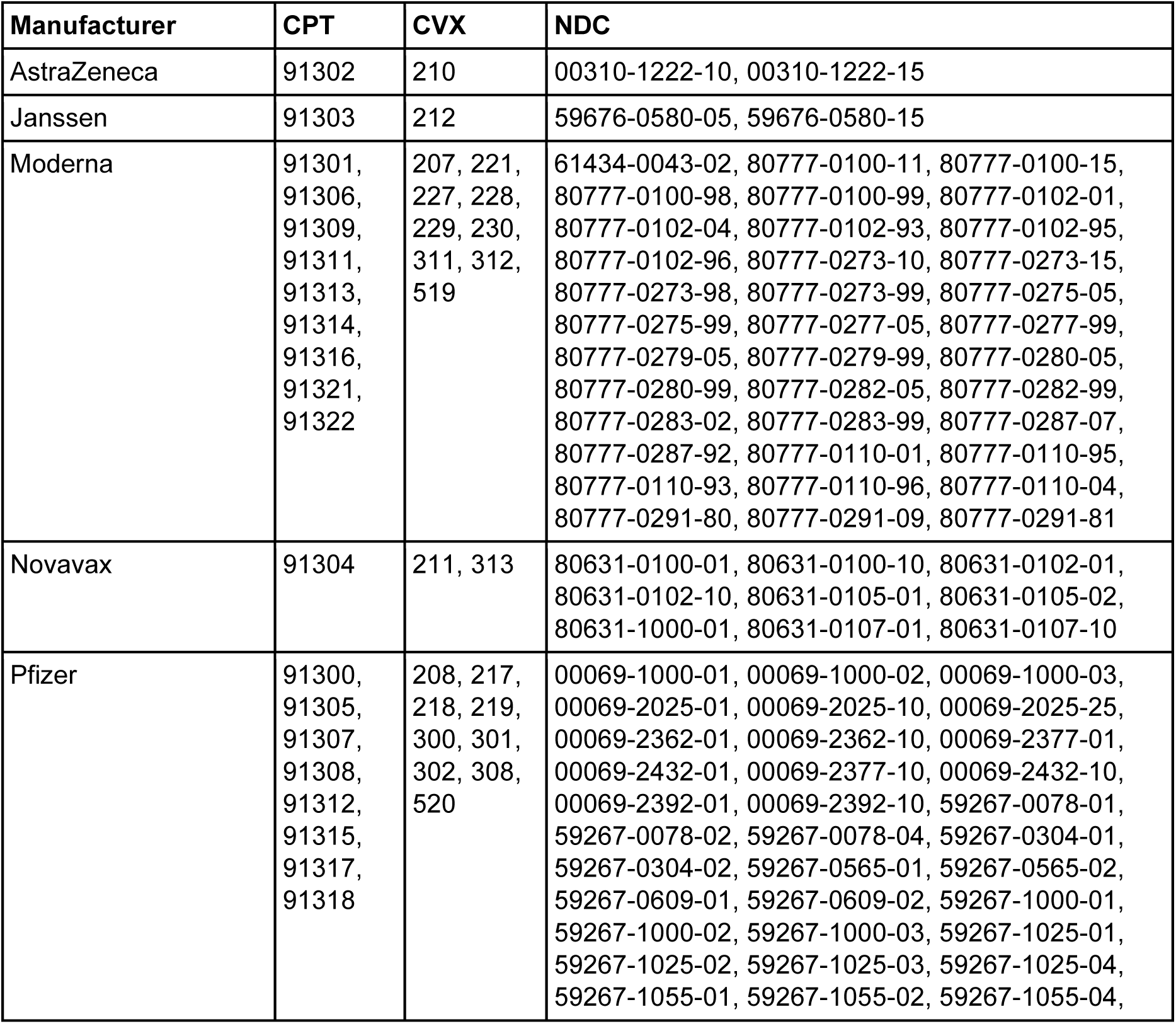
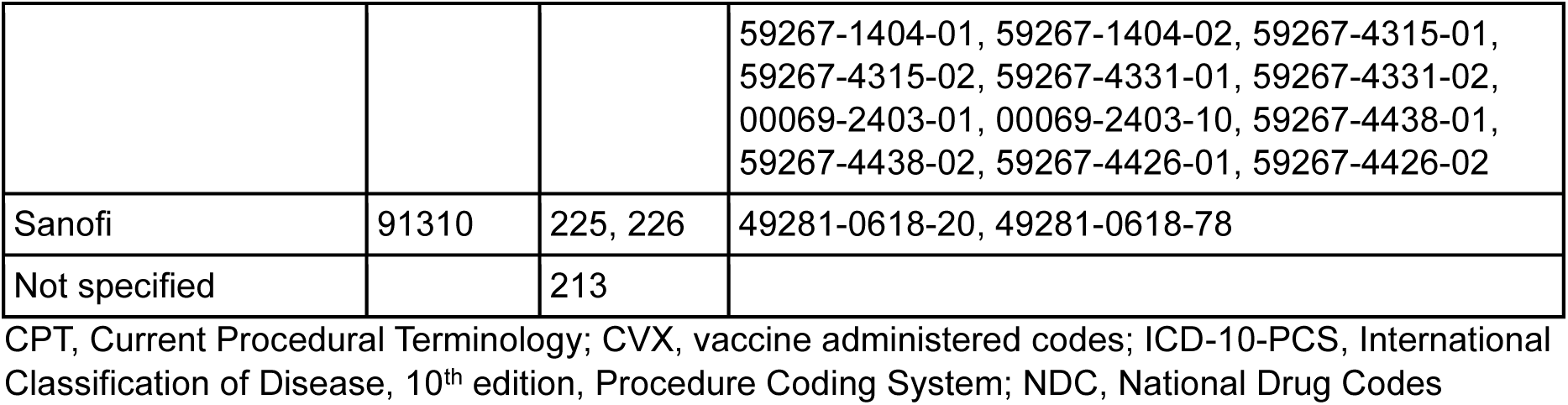
List of CVX, CPT, and NDC codes used to identify COVID-19 vaccines other than formulations used to define the index event.

**Supplementary Table 3.** Codes used to identify COVID-19 diagnosis or treatment.

| Category | Code Type | Codes for COVID-related medical encounters |
| --- | --- | --- |
| COVID-19 treatment | Generic drug name | abatacept, anakinra, bamlanivimab, baricitinib, bebtelovimab, betamethasone, budesonide, bupivacaine/dexamethasone, casirivimab/imdevimab, cortisone acetate, deflazacort, dexamethasone, hydrocortisone, infliximab, methylprednisolone, molnupiravir, nirmatrelvir/ritonavir, prednisolone, prednisone, remdesivir, sotrovimab, tocilizumab, triamcinolone, vilobelimab |
| COVID-19 treatment | HCPCS | M0245, M0246, Q0245, M0222, M0223, Q0222, M0240, M0241, M0243, M0244, Q0240, Q0243, Q0244, J0248, M0247, M0248, Q0247, J3262, M0249, M0250, Q0249, J0129, J1745, Q5102, Q5103, Q5104, Q5109, Q5121, C9469, J0702, J1020, J1030, J1040, J1094, J1100, J1700, J1710, J1720, J2650, J2920, J2930, J3300, J3301, J3302, J3303, J3304, J7509, J7510, J8540, Q9993 |
| COVID-19 treatment | CPT | 94002, 94003, 94004, 94660, 94662, 33946, 33947, 33948, 33949, 33951, 33952, 33953, 33954, 33955, 33956, 33957, 33958, 33959, 33962, 33963, 33964, 33965, 33966, 33969, 33984, 33985, 33986 |
| COVID-19 treatment | DRG | 207, 208 |
| COVID-19 treatment | ICD-10-PCS | XW033F6, XW043F6, XW033E5, XW043E5, XW033H5, XW043H5, 5A09357, 5A09358, 5A09359, 5A0935A, 5A0935B, 5A0935Z, 5A09457, 5A09458, 5A09459, 5A0945A, 5A0945B, 5A0945Z, 5A09557, 5A09558, 5A09559, 5A0955A, 5A0955B, 5A0955Z, 5A09B5K, 5A09C5K, 5A09D5K, 5A19054, 5A1935Z, 5A1945Z, 5A1955Z, 5A1522F, 5A1522G, 5A1522H, 5A15A2F, 5A15A2G, 5A15A2H |
| COVID-19 diagnosis | ICD-10-CM | J1282, U071 |
| COVID-19 diagnosis | SNOMED | 1119302008, 119731000146105, 119741000146102, 119751000146104, 119981000146107, 1240411000000107, 1240521000000100, 1240531000000103, 1240541000000107, 1240561000000108, 1240581000000104, 674814021000119106, 840533007, 840534001, 840536004, 840539006, 866151004, |
|  |  | 866152006, 870588003, 870589006, 870590002, 870591003,<br>871562009 |

CPT, Current Procedural Terminology; DRG, Diagnosis-Related Group; HCPCS, Healthcare Common Procedure Coding System; ICD-10-PCS, International Classification of Disease, 10<sup>th</sup> edition, Procedure Coding System; ICD-10-CM, International Classification of Disease, 10<sup>th</sup> edition, Clinical Modification, SNOMED, Systemized Nomenclature of Medicine

**Supplementary Table 4.** Baseline Characteristics (Pre-weighting and Post-weighting, and SMDs) – mRNA-1283 ≥65 Years.

Pre-weighting N: 233,072 vaccinated / 233,072 comparator | Post-weighting N: 230,778 vaccinated / 232,727 comparator
|  | Pre-weighting |  |  | Post-weighting (IPTW-adjusted) |  |  |
| --- | --- | --- | --- | --- | --- | --- |
|  | Vaccinated | Comparator | SMD | Vaccinated | Comparator | SMD |
| **Number of Patients** | 233,072 | 233,072 |  | 230,778 | 232,727 |  |
| **Age; Mean (SD)** | 76 (6.4) | 76 (6.4) |  | 76 (6.3) | 76 (6.4) | 0.0020 |
| **Age Categories; n (%)** |  |  |  |  |  | 0.0019 |
| 65-69 | 42,356 (18.2%) | 42,356 (18.2%) |  | 41,980 (18.2%) | 42,282 (18.2%) |  |
| 70-74 | 64,580 (27.7%) | 64,580 (27.7%) |  | 63,734 (27.6%) | 64,374 (27.7%) |  |
| 75-79 | 59,698 (25.6%) | 59,698 (25.6%) |  | 58,952 (25.5%) | 59,561 (25.6%) |  |
| 80-84 | 37,799 (16.2%) | 37,799 (16.2%) |  | 37,473 (16.2%) | 37,773 (16.2%) |  |
| 85-89 | 28,639 (12.3%) | 28,639 (12.3%) |  | 28,639 (12.4%) | 28,737 (12.3%) |  |
| **Sex; n (%)** |  |  |  |  |  | 0.0003 |
| Male | 102,160 (43.8%) | 102,160 (43.8%) |  | 101,199 (43.9%) | 102,090 (43.9%) |  |
| **Race; n (%)** |  |  |  |  |  | 0.0006 |
| Black | 13,459 (5.8%) | 13,459 (5.8%) |  | 13,339 (5.8%) | 13,427 (5.8%) |  |
| White | 150,841 (64.7%) | 150,841 (64.7%) |  | 149,381 (64.7%) | 150,704 (64.8%) |  |
| Other | 31,923 (13.7%) | 31,923 (13.7%) |  | 31,628 (13.7%) | 31,860 (13.7%) |  |
| Unknown | 36,849 (15.8%) | 36,849 (15.8%) |  | 36,431 (15.8%) | 36,737 (15.8%) |  |
| **Ethnicity; n (%)** |  |  |  |  |  | 0.0006 |
| Hispanic | 4,107 (1.8%) | 4,107 (1.8%) |  | 4,091 (1.8%) | 4,110 (1.8%) |  |
| Non-Hispanic | 138,819 (59.6%) | 138,819 (59.6%) |  | 137,570 (59.6%) | 138,664 (59.6%) |  |
| Unknown | 90,146 (38.7%) | 90,146 (38.7%) |  | 89,117 (38.6%) | 89,953 (38.7%) |  |
| **Insurance Type; n (%)** |  |  | 0.1484 |  |  | 0.0081 |
| Commercial | 14,301 (6.1%) | 12,951 (5.6%) |  | 13,561 (5.9%) | 13,624 (5.9%) |  |
| Medicaid | 1,529 (0.7%) | 2,989 (1.3%) |  | 2,224 (1.0%) | 2,275 (1.0%) |  |
| Medicare Advantage | 193,885 (83.2%) | 186,697 (80.1%) |  | 188,605 (81.7%) | 189,925 (81.6%) |  |
| Medicare FFS | 19,677 (8.4%) | 22,255 (9.5%) |  | 20,786 (9.0%) | 20,983 (9.0%) |  |
| Unknown | 3,680 (1.6%) | 8,180 (3.5%) |  | 5,602 (2.4%) | 5,919 (2.5%) |  |
| **Region; n (%)** |  |  |  |  |  | 0.0016 |
| Midwest | 43,668 (18.7%) | 43,668 (18.7%) |  | 43,372 (18.8%) | 43,665 (18.8%) |  |
| Northeast | 63,554 (27.3%) | 63,554 (27.3%) |  | 62,998 (27.3%) | 63,451 (27.3%) |  |
| South | 68,098 (29.2%) | 68,098 (29.2%) |  | 67,358 (29.2%) | 67,952 (29.2%) |  |
| West | 57,751 (24.8%) | 57,751 (24.8%) |  | 57,049 (24.7%) | 57,658 (24.8%) |  |
| Unknown | 1 (0.0%) | 1 (0.0%) |  | 1 (0.0%) | 1 (0.0%) |  |
| **Month of Index Date; n (%)** |  |  |  |  |  | 0.0100 |
| 2025-08 | 59 (0.0%) | 59 (0.0%) |  | 60 (0.0%) | 56 (0.0%) |  |
| 2025-09 | 89,220 (38.3%) | 89,220 (38.3%) |  | 88,353 (38.3%) | 88,763 (38.1%) |  |
| 2025-10 | 98,016 (42.1%) | 98,016 (42.1%) |  | 97,082 (42.1%) | 97,723 (42.0%) |  |
| 2025-11 | 34,095 (14.6%) | 34,095 (14.6%) |  | 33,738 (14.6%) | 34,303 (14.7%) |  |
| 2025-12 | 10,061 (4.3%) | 10,061 (4.3%) |  | 9,953 (4.3%) | 10,181 (4.4%) |  |
| 2026-01 | 1,621 (0.7%) | 1,621 (0.7%) |  | 1,593 (0.7%) | 1,700 (0.7%) |  |
| **Number of OP Visits; Mean (SD)** | 2.4 (5.8) | 2.7 (6.5) | 0.0422 | 2.6 (6.1) | 2.6 (6.2) | 0.0013 |
| **Number of Hospitalizations; Mean (SD)** | 0.1 (0.4) | 0.2 (0.5) | 0.0853 | 0.1 (0.5) | 0.1 (0.5) | 0.0014 |
| **Baseline Clinical Comorbidities; n (%)** |  |  |  |  |  |  |
| Cancer | 38,136 (16.4%) | 37,120 (15.9%) | 0.0118 | 37,307 (16.2%) | 37,580 (16.1%) | 0.0005 |
| Cerebrovascular disease | 28,531 (12.2%) | 33,925 (14.6%) | 0.0680 | 30,834 (13.4%) | 31,213 (13.4%) | 0.0015 |
| Chronic kidney disease | 56,270 (24.1%) | 64,490 (27.7%) | 0.0806 | 59,766 (25.9%) | 60,402 (26.0%) | 0.0013 |
| Chronic lung diseases | 37,033 (15.9%) | 45,180 (19.4%) | 0.0918 | 40,651 (17.6%) | 41,135 (17.7%) | 0.0016 |
| Chronic liver diseases | 4,037 (1.7%) | 4,897 (2.1%) | 0.0269 | 4,415 (1.9%) | 4,469 (1.9%) | 0.0005 |
| Cystic fibrosis | 35 (0.0%) | 29 (0.0%) | 0.0022 | 32 (0.0%) | 32 (0.0%) | 0.0003 |
| Diabetes mellitus, type 1 and type 2 | 71,558 (30.7%) | 83,439 (35.8%) | 0.1084 | 76,794 (33.3%) | 77,576 (33.3%) | 0.0012 |
| Disabilities | 13,501 (5.8%) | 13,087 (5.6%) | 0.0077 | 13,170 (5.7%) | 13,270 (5.7%) | 0.0002 |
| Heart conditions | 73,954 (31.7%) | 82,284 (35.3%) | 0.0758 | 77,366 (33.5%) | 78,111 (33.6%) | 0.0008 |
| HIV | 770 (0.3%) | 660 (0.3%) | 0.0085 | 709 (0.3%) | 712 (0.3%) | 0.0002 |
| Mental health disorders | 49,968 (21.4%) | 58,509 (25.1%) | 0.0868 | 53,716 (23.3%) | 54,260 (23.3%) | 0.0009 |
| Neurologic conditions | 15,778 (6.8%) | 23,421 (10.0%) | 0.1184 | 19,377 (8.4%) | 19,648 (8.4%) | 0.0017 |
| Obesity | 61,980 (26.6%) | 68,204 (29.3%) | 0.0595 | 64,559 (28.0%) | 65,161 (28.0%) | 0.0005 |
| Primary Immunodeficiencies | 5,919 (2.5%) | 6,755 (2.9%) | 0.0221 | 6,260 (2.7%) | 6,336 (2.7%) | 0.0006 |
| Physical inactivity | 398 (0.2%) | 489 (0.2%) | 0.0090 | 447 (0.2%) | 446 (0.2%) | 0.0005 |
| Smoking, current and former | 48,815 (20.9%) | 55,045 (23.6%) | 0.0643 | 51,432 (22.3%) | 51,958 (22.3%) | 0.0009 |
| Solid organ or hematopoietic cell transplantation | 722 (0.3%) | 807 (0.3%) | 0.0064 | 762 (0.3%) | 770 (0.3%) | 0.0001 |
| Tuberculosis | 86 (0.0%) | 112 (0.0%) | 0.0054 | 100 (0.0%) | 100 (0.0%) | 0.0002 |
| Use of immunosuppressive medications | 18,523 (7.9%) | 16,249 (7.0%) | 0.0371 | 17,274 (7.5%) | 17,370 (7.5%) | 0.0008 |
| Asthma | 22,162 (9.5%) | 21,635 (9.3%) | 0.0077 | 21,661 (9.4%) | 21,842 (9.4%) |  |
| **Time since last COVID monovalent vaccination** |  |  | 0.2022 |  |  | 0.0754 |
| <= 90 days | 1,018 (0.4%) | 7,135 (3.1%) |  | 2,057 (0.9%) | 4,080 (1.8%) |  |
| 91-180 days | 19,789 (8.5%) | 20,435 (8.8%) |  | 20,055 (8.7%) | 20,108 (8.6%) |  |
| 180+ days | 176,631 (75.8%) | 169,811 (72.9%) |  | 172,973 (75.0%) | 173,064 (74.4%) |  |
| Not reported | 35,634 (15.3%) | 35,691 (15.3%) |  | 35,692 (15.5%) | 35,475 (15.2%) |  |
| **Time since last COVID infection** |  |  | 0.0340 |  |  | 0.0025 |
| <= 120 days | 478 (0.2%) | 474 (0.2%) |  | 472 (0.2%) | 476 (0.2%) |  |
| 121-180 days | 769 (0.3%) | 695 (0.3%) |  | 725 (0.3%) | 728 (0.3%) |  |
| 180+ days | 6,638 (2.8%) | 5,410 (2.3%) |  | 5,945 (2.6%) | 5,913 (2.5%) |  |
| Not reported | 225,187 (96.6%) | 226,493 (97.2%) |  | 223,636 (96.9%) | 225,609 (96.9%) |  |
| **Evidence of COVID vaccination with prior season's vaccine during the pre-index period** | 197,307 (84.7%) | 197,307 (84.7%) |  | 194,979 (84.5%) | 197,158 (84.7%) | 0.0063 |
| **Evidence of influenza vaccination during the pre-index period** | 210,509 (90.3%) | 191,766 (82.3%) | 0.2355 | 199,037 (86.2%) | 200,610 (86.2%) | 0.0013 |
| **Diagnosis of influenza during pre-index period** | 4,219 (1.8%) | 4,624 (2.0%) | 0.0127 | 4,411 (1.9%) | 4,439 (1.9%) | 0.0003 |
| **Week of last healthcare activity before start of intake** |  |  |  |  |  | 0.0055 |
| 0-4 weeks | 214,304 (91.9%) | 214,304 (91.9%) |  | 212,213 (92.0%) | 214,069 (92.0%) |  |
| 5-26 weeks | 18,480 (7.9%) | 18,480 (7.9%) |  | 18,281 (7.9%) | 18,373 (7.9%) |  |
| 27-52 weeks | 217 (0.1%) | 217 (0.1%) |  | 212 (0.1%) | 214 (0.1%) |  |
| > 52 weeks or unknown | 71 (0.0%) | 71 (0.0%) |  | 73 (0.0%) | 71 (0.0%) |  |

SMD = standardized mean difference. Values with absolute SMD >0.10 indicate covariate imbalance. Post-weighting values (IPTW-adjusted) are used in Table 1 characteristics; pre-weighting patient counts are used in all text. Vaccine counts (n) in the comorbidity rows reflect presence of the condition (=1).

**Supplementary Table 5.** Baseline Characteristics (Pre-weighting and Post-weighting, and SMDs) – BNT162b2 ≥65 Years.

Pre-weighting N: 422,610 vaccinated / 422,610 comparator | Post-weighting N: 418,590 vaccinated / 422,109 comparator
|  | Pre-weighting |  |  | Post-weighting (IPTW-adjusted) |  |  |
| --- | --- | --- | --- | --- | --- | --- |
|  | Vaccinated | Comparator | SMD | Vaccinated | Comparator | SMD |
| **Number of Patients** | 422,610 | 422,610 |  | 418,590 | 422,109 |  |
| **Age; Mean (SD)** | 76 (6.4) | 76 (6.4) |  | 76 (6.4) | 76 (6.4) | 0.0016 |
| **Age Categories; n (%)** |  |  |  |  |  | 0.0015 |
| 65-69 | 80,765 (19.1%) | 80,765 (19.1%) |  | 79,999 (19.1%) | 80,572 (19.1%) |  |
| 70-74 | 113,856 (26.9%) | 113,856 (26.9%) |  | 112,558 (26.9%) | 113,637 (26.9%) |  |
| 75-79 | 105,977 (25.1%) | 105,977 (25.1%) |  | 104,753 (25.0%) | 105,779 (25.1%) |  |
| 80-84 | 69,025 (16.3%) | 69,025 (16.3%) |  | 68,449 (16.4%) | 69,006 (16.3%) |  |
| 85-89 | 52,987 (12.5%) | 52,987 (12.5%) |  | 52,831 (12.6%) | 53,116 (12.6%) |  |
| **Sex; n (%)** |  |  |  |  |  |  |
| Male | 184,699 (43.7%) | 184,699 (43.7%) |  | 183,012 (43.7%) | 184,548 (43.7%) |  |
| **Race; n (%)** |  |  |  |  |  | 0.0010 |
| Black | 32,048 (7.6%) | 32,048 (7.6%) |  | 31,815 (7.6%) | 31,980 (7.6%) |  |
| White | 266,619 (63.1%) | 266,619 (63.1%) |  | 264,017 (63.1%) | 266,412 (63.1%) |  |
| Other | 59,138 (14.0%) | 59,138 (14.0%) |  | 58,635 (14.0%) | 59,097 (14.0%) |  |
| Unknown | 64,805 (15.3%) | 64,805 (15.3%) |  | 64,123 (15.3%) | 64,620 (15.3%) |  |
| **Ethnicity; n (%)** |  |  |  |  |  | 0.0008 |
| Hispanic | 8,934 (2.1%) | 8,934 (2.1%) |  | 8,907 (2.1%) | 8,933 (2.1%) |  |
| Non-Hispanic | 250,383 (59.2%) | 250,383 (59.2%) |  | 248,093 (59.3%) | 250,161 (59.3%) |  |
| Unknown | 163,293 (38.6%) | 163,293 (38.6%) |  | 161,590 (38.6%) | 163,015 (38.6%) |  |
| **Insurance Type; n (%)** |  |  | 0.1060 |  |  | 0.0041 |
| Commercial | 27,486 (6.5%) | 24,097 (5.7%) |  | 25,627 (6.1%) | 25,727 (6.1%) |  |
| Medicaid | 2,643 (0.6%) | 5,310 (1.3%) |  | 3,797 (0.9%) | 3,987 (0.9%) |  |
| Medicare Advantage | 347,445 (82.2%) | 338,200 (80.0%) |  | 339,763 (81.2%) | 342,376 (81.1%) |  |
| Medicare FFS | 33,485 (7.9%) | 38,916 (9.2%) |  | 35,853 (8.6%) | 36,248 (8.6%) |  |
| Unknown | 11,551 (2.7%) | 16,087 (3.8%) |  | 13,549 (3.2%) | 13,771 (3.3%) |  |
| **Region; n (%)** |  |  |  |  |  | 0.0014 |
| Midwest | 76,230 (18.0%) | 76,230 (18.0%) |  | 75,696 (18.1%) | 76,219 (18.1%) |  |
| Northeast | 92,481 (21.9%) | 92,481 (21.9%) |  | 91,658 (21.9%) | 92,383 (21.9%) |  |
| South | 153,879 (36.4%) | 153,879 (36.4%) |  | 152,450 (36.4%) | 153,620 (36.4%) |  |
| West | 100,014 (23.7%) | 100,014 (23.7%) |  | 98,781 (23.6%) | 99,882 (23.7%) |  |
| Unknown | 6 (0.0%) | 6 (0.0%) |  | 5 (0.0%) | 6 (0.0%) |  |
| **Month of Index Date; n (%)** |  |  |  |  |  | 0.0078 |
| 2025-08 | 153 (0.0%) | 153 (0.0%) |  | 158 (0.0%) | 154 (0.0%) |  |
| 2025-09 | 162,276 (38.4%) | 162,276 (38.4%) |  | 160,945 (38.4%) | 161,372 (38.2%) |  |
| 2025-10 | 175,617 (41.6%) | 175,617 (41.6%) |  | 173,781 (41.5%) | 175,113 (41.5%) |  |
| 2025-11 | 61,306 (14.5%) | 61,306 (14.5%) |  | 60,671 (14.5%) | 61,733 (14.6%) |  |
| 2025-12 | 20,375 (4.8%) | 20,375 (4.8%) |  | 20,227 (4.8%) | 20,712 (4.9%) |  |
| 2026-01 | 2,883 (0.7%) | 2,883 (0.7%) |  | 2,808 (0.7%) | 3,025 (0.7%) |  |
| **Number of OP Visits; Mean (SD)** | 2.5 (6.0) | 2.7 (6.5) | 0.0319 | 2.6 (6.3) | 2.6 (6.2) | 0.0012 |
| **Number of Hospitalizations; Mean (SD)** | 0.1 (0.4) | 0.2 (0.5) | 0.0805 | 0.1 (0.5) | 0.1 (0.5) | 0.0025 |
| **Baseline Clinical Comorbidities; n (%)** |  |  |  |  |  |  |
| Cancer | 68,241 (16.1%) | 66,687 (15.8%) | 0.0100 | 66,871 (16.0%) | 67,413 (16.0%) | 0.0001 |
| Cerebrovascular disease | 53,845 (12.7%) | 62,386 (14.8%) | 0.0587 | 57,470 (13.7%) | 58,127 (13.8%) | 0.0012 |
| Chronic kidney disease | 113,142 (26.8%) | 120,988 (28.6%) | 0.0415 | 116,030 (27.7%) | 117,075 (27.7%) | 0.0004 |
| Chronic lung diseases | 72,696 (17.2%) | 84,061 (19.9%) | 0.0692 | 77,618 (18.5%) | 78,430 (18.6%) | 0.0010 |
| Chronic liver diseases | 7,620 (1.8%) | 8,840 (2.1%) | 0.0209 | 8,117 (1.9%) | 8,221 (1.9%) | 0.0006 |
| Cystic fibrosis | 63 (0.0%) | 59 (0.0%) | 0.0008 | 58 (0.0%) | 59 (0.0%) | 0.0001 |
| Diabetes mellitus, type 1 and type 2 | 138,839 (32.9%) | 154,363 (36.5%) | 0.0772 | 145,460 (34.8%) | 146,758 (34.8%) | 0.0004 |
| Disabilities | 24,038 (5.7%) | 23,492 (5.6%) | 0.0056 | 23,522 (5.6%) | 23,724 (5.6%) |  |
| Heart conditions | 140,038 (33.1%) | 151,944 (36.0%) | 0.0593 | 144,610 (34.5%) | 145,996 (34.6%) | 0.0009 |
| HIV | 1,565 (0.4%) | 1,221 (0.3%) | 0.0142 | 1,391 (0.3%) | 1,386 (0.3%) | 0.0007 |
| Mental health disorders | 94,790 (22.4%) | 106,279 (25.1%) | 0.0639 | 99,616 (23.8%) | 100,576 (23.8%) | 0.0007 |
| Neurologic conditions | 30,835 (7.3%) | 42,859 (10.1%) | 0.1010 | 36,429 (8.7%) | 36,906 (8.7%) | 0.0014 |
| Obesity | 119,616 (28.3%) | 125,481 (29.7%) | 0.0306 | 121,633 (29.1%) | 122,659 (29.1%) |  |
| Primary Immunodeficiencies | 12,254 (2.9%) | 12,638 (3.0%) | 0.0054 | 12,320 (2.9%) | 12,435 (2.9%) | 0.0002 |
| Physical inactivity | 813 (0.2%) | 930 (0.2%) | 0.0061 | 862 (0.2%) | 871 (0.2%) | 0.0001 |
| Smoking, current and former | 91,814 (21.7%) | 101,551 (24.0%) | 0.0549 | 95,673 (22.9%) | 96,642 (22.9%) | 0.0009 |
| Solid organ or hematopoietic cell transplantation | 1,455 (0.3%) | 1,515 (0.4%) | 0.0024 | 1,475 (0.4%) | 1,489 (0.4%) | 0.0001 |
| Tuberculosis | 157 (0.0%) | 229 (0.1%) | 0.0080 | 193 (0.0%) | 196 (0.0%) | 0.0001 |
| Use of immunosuppressive medications | 33,008 (7.8%) | 29,403 (7.0%) | 0.0326 | 30,977 (7.4%) | 31,164 (7.4%) | 0.0007 |
| Asthma | 40,951 (9.7%) | 39,453 (9.3%) | 0.0121 | 39,825 (9.5%) | 40,143 (9.5%) | 0.0001 |
| **Time since last COVID monovalent vaccination** |  |  | 0.1962 |  |  | 0.0808 |
| <= 90 days | 1,623 (0.4%) | 11,724 (2.8%) |  | 3,032 (0.7%) | 6,679 (1.6%) |  |
| 91-180 days | 28,091 (6.6%) | 30,866 (7.3%) |  | 29,227 (7.0%) | 29,396 (7.0%) |  |
| 180+ days | 322,225 (76.2%) | 309,246 (73.2%) |  | 315,517 (75.4%) | 315,591 (74.8%) |  |
| Not reported | 70,671 (16.7%) | 70,774 (16.7%) |  | 70,815 (16.9%) | 70,444 (16.7%) |  |
| **Time since last COVID infection** |  |  | 0.0234 |  |  | 0.0013 |
| <= 120 days | 874 (0.2%) | 944 (0.2%) |  | 891 (0.2%) | 901 (0.2%) |  |
| 121-180 days | 1,355 (0.3%) | 1,222 (0.3%) |  | 1,268 (0.3%) | 1,278 (0.3%) |  |
| 180+ days | 11,129 (2.6%) | 9,645 (2.3%) |  | 10,252 (2.4%) | 10,259 (2.4%) |  |
| Not reported | 409,252 (96.8%) | 410,799 (97.2%) |  | 406,180 (97.0%) | 409,671 (97.1%) |  |
| **Evidence of COVID vaccination with prior season's vaccine during the pre-index period** | 351,664 (83.2%) | 351,664 (83.2%) |  | 347,550 (83.0%) | 351,456 (83.3%) | 0.0062 |
| **Evidence of influenza vaccination during the pre-index period** | 379,437 (89.8%) | 343,666 (81.3%) | 0.2425 | 357,822 (85.5%) | 360,756 (85.5%) | 0.0005 |
| **Diagnosis of influenza during pre-index period** | 8,117 (1.9%) | 8,502 (2.0%) | 0.0066 | 8,238 (2.0%) | 8,313 (2.0%) | 0.0001 |
| **Week of last healthcare activity before start of intake** |  |  |  |  |  | 0.0015 |
| 0-4 weeks | 389,493 (92.2%) | 389,493 (92.2%) |  | 385,774 (92.2%) | 389,201 (92.2%) |  |
| 5-26 weeks | 32,527 (7.7%) | 32,527 (7.7%) |  | 32,236 (7.7%) | 32,334 (7.7%) |  |
| 27-52 weeks | 444 (0.1%) | 444 (0.1%) |  | 437 (0.1%) | 432 (0.1%) |  |
| > 52 weeks or unknown | 146 (0.0%) | 146 (0.0%) |  | 143 (0.0%) | 142 (0.0%) |  |

SMD = standardized mean difference. Values with absolute SMD >0.10 indicate covariate imbalance. Post-weighting values (IPTW-adjusted) are used in Table 1 characteristics; pre-weighting patient counts are used in all text. Vaccine counts (n) in the comorbidity rows reflect presence of the condition (=1).

**Supplementary Table 6.** Baseline Characteristics (Pre-weighting and Post-weighting, and SMDs) – mRNA-1283 ≥75 Years.

Pre-weighting N: 126,136 vaccinated / 126,136 comparator | Post-weighting N: 124,862 vaccinated / 125,966 comparator
|  | Pre-weighting |  |  | Post-weighting (IPTW-adjusted) |  |  |
| --- | --- | --- | --- | --- | --- | --- |
|  | Vaccinated | Comparator | SMD | Vaccinated | Comparator | SMD |
| **Number of Patients** | 126,136 | 126,136 |  | 124,862 | 125,966 |  |
| **Age; Mean (SD)** | 81 (4.2) | 81 (4.2) |  | 81 (4.2) | 81 (4.2) | 0.0053 |
| **Age Categories; n (%)** |  |  |  |  |  | 0.0028 |
| 75-79 | 59,698 (47.3%) | 59,698 (47.3%) |  | 58,866 (47.1%) | 59,518 (47.2%) |  |
| 80-84 | 37,799 (30.0%) | 37,799 (30.0%) |  | 37,411 (30.0%) | 37,741 (30.0%) |  |
| 85-89 | 28,639 (22.7%) | 28,639 (22.7%) |  | 28,585 (22.9%) | 28,707 (22.8%) |  |
| **Sex; n (%)** |  |  |  |  |  | 0.0008 |
| Male | 54,925 (43.5%) | 54,925 (43.5%) |  | 54,362 (43.5%) | 54,895 (43.6%) |  |
| **Race; n (%)** |  |  |  |  |  | 0.0009 |
| Black | 6,447 (5.1%) | 6,447 (5.1%) |  | 6,384 (5.1%) | 6,426 (5.1%) |  |
| White | 83,851 (66.5%) | 83,851 (66.5%) |  | 83,021 (66.5%) | 83,795 (66.5%) |  |
| Other | 17,225 (13.7%) | 17,225 (13.7%) |  | 17,070 (13.7%) | 17,196 (13.7%) |  |
| Unknown | 18,613 (14.8%) | 18,613 (14.8%) |  | 18,386 (14.7%) | 18,548 (14.7%) |  |
| **Ethnicity; n (%)** |  |  |  |  |  | 0.0010 |
| Hispanic | 1,915 (1.5%) | 1,915 (1.5%) |  | 1,911 (1.5%) | 1,920 (1.5%) |  |
| Non-Hispanic | 75,650 (60.0%) | 75,650 (60.0%) |  | 74,927 (60.0%) | 75,564 (60.0%) |  |
| Unknown | 48,571 (38.5%) | 48,571 (38.5%) |  | 48,024 (38.5%) | 48,482 (38.5%) |  |
| **Insurance Type; n (%)** |  |  | 0.1394 |  |  | 0.0076 |
| Commercial | 1,751 (1.4%) | 1,762 (1.4%) |  | 1,751 (1.4%) | 1,761 (1.4%) |  |
| Medicaid | 509 (0.4%) | 1,153 (0.9%) |  | 792 (0.6%) | 835 (0.7%) |  |
| Medicare Advantage | 111,260 (88.2%) | 106,953 (84.8%) |  | 108,133 (86.6%) | 108,916 (86.5%) |  |
| Medicare FFS | 11,059 (8.8%) | 12,755 (10.1%) |  | 11,800 (9.5%) | 11,925 (9.5%) |  |
| Unknown | 1,557 (1.2%) | 3,513 (2.8%) |  | 2,387 (1.9%) | 2,530 (2.0%) |  |
| **Region; n (%)** |  |  |  |  |  | 0.0020 |
| Midwest | 22,761 (18.0%) | 22,761 (18.0%) |  | 22,625 (18.1%) | 22,776 (18.1%) |  |
| Northeast | 34,332 (27.2%) | 34,332 (27.2%) |  | 34,033 (27.3%) | 34,288 (27.2%) |  |
| South | 36,852 (29.2%) | 36,852 (29.2%) |  | 36,458 (29.2%) | 36,781 (29.2%) |  |
| West | 32,191 (25.5%) | 32,191 (25.5%) |  | 31,746 (25.4%) | 32,121 (25.5%) |  |
| Unknown |  |  |  |  |  |  |
| **Month of Index Date; n (%)** |  |  |  |  |  | 0.0056 |
| 2025-08 | 22 (0.0%) | 22 (0.0%) |  | 23 (0.0%) | 21 (0.0%) |  |
| 2025-09 | 48,020 (38.1%) | 48,020 (38.1%) |  | 47,462 (38.0%) | 47,806 (38.0%) |  |
| 2025-10 | 54,351 (43.1%) | 54,351 (43.1%) |  | 53,892 (43.2%) | 54,220 (43.0%) |  |
| 2025-11 | 17,952 (14.2%) | 17,952 (14.2%) |  | 17,767 (14.2%) | 18,031 (14.3%) |  |
| 2025-12 | 5,003 (4.0%) | 5,003 (4.0%) |  | 4,933 (4.0%) | 5,071 (4.0%) |  |
| 2026-01 | 788 (0.6%) | 788 (0.6%) |  | 786 (0.6%) | 817 (0.6%) |  |
| **Number of OP Visits; Mean (SD)** | 2.5 (5.9) | 2.8 (6.5) | 0.0392 | 2.7 (6.2) | 2.7 (6.2) | 0.0012 |
| **Number of Hospitalizations; Mean (SD)** | 0.1 (0.4) | 0.2 (0.5) | 0.0888 | 0.2 (0.5) | 0.2 (0.5) | 0.0016 |
| **Baseline Clinical Comorbidities; n (%)** |  |  |  |  |  |  |
| Cancer | 23,367 (18.5%) | 22,506 (17.8%) | 0.0177 | 22,755 (18.2%) | 22,921 (18.2%) | 0.0007 |
| Cerebrovascular disease | 19,212 (15.2%) | 22,551 (17.9%) | 0.0713 | 20,617 (16.5%) | 20,872 (16.6%) | 0.0016 |
| Chronic kidney disease | 38,566 (30.6%) | 43,528 (34.5%) | 0.0840 | 40,618 (32.5%) | 41,051 (32.6%) | 0.0012 |
| Chronic lung diseases | 23,707 (18.8%) | 28,160 (22.3%) | 0.0874 | 25,680 (20.6%) | 25,959 (20.6%) | 0.0010 |
| Chronic liver diseases | 1,895 (1.5%) | 2,273 (1.8%) | 0.0235 | 2,063 (1.7%) | 2,084 (1.7%) | 0.0002 |
| Cystic fibrosis | 19 (0.0%) | 11 (0.0%) | 0.0058 | 15 (0.0%) | 14 (0.0%) | 0.0009 |
| Diabetes mellitus, type 1 and type 2 | 39,059 (31.0%) | 45,713 (36.2%) | 0.1119 | 41,948 (33.6%) | 42,415 (33.7%) | 0.0016 |
| Disabilities | 6,836 (5.4%) | 6,607 (5.2%) | 0.0081 | 6,655 (5.3%) | 6,705 (5.3%) | 0.0003 |
| Heart conditions | 47,188 (37.4%) | 52,000 (41.2%) | 0.0782 | 49,110 (39.3%) | 49,582 (39.4%) | 0.0006 |
| HIV | 188 (0.1%) | 166 (0.1%) | 0.0047 | 174 (0.1%) | 174 (0.1%) | 0.0004 |
| Mental health disorders | 26,615 (21.1%) | 31,381 (24.9%) | 0.0899 | 28,716 (23.0%) | 29,013 (23.0%) | 0.0008 |
| Neurologic conditions | 12,918 (10.2%) | 18,767 (14.9%) | 0.1403 | 15,673 (12.6%) | 15,877 (12.6%) | 0.0016 |
| Obesity | 28,607 (22.7%) | 31,682 (25.1%) | 0.0572 | 29,868 (23.9%) | 30,163 (23.9%) | 0.0006 |
| Primary Immunodeficiencies | 3,194 (2.5%) | 3,677 (2.9%) | 0.0235 | 3,401 (2.7%) | 3,439 (2.7%) | 0.0004 |
| Physical inactivity | 242 (0.2%) | 281 (0.2%) | 0.0068 | 263 (0.2%) | 263 (0.2%) | 0.0005 |
| Smoking, current and former | 26,601 (21.1%) | 28,830 (22.9%) | 0.0427 | 27,458 (22.0%) | 27,729 (22.0%) | 0.0005 |
| Solid organ or hematopoietic cell transplantation | 263 (0.2%) | 265 (0.2%) | 0.0003 | 266 (0.2%) | 268 (0.2%) | 0.0002 |
| Tuberculosis | 50 (0.0%) | 75 (0.1%) | 0.0089 | 63 (0.1%) | 64 (0.1%) | 0.0001 |
| Use of immunosuppressive medications | 10,211 (8.1%) | 8,734 (6.9%) | 0.0444 | 9,407 (7.5%) | 9,461 (7.5%) | 0.0009 |
| Asthma | 11,254 (8.9%) | 11,132 (8.8%) | 0.0034 | 11,059 (8.9%) | 11,161 (8.9%) | 0.0001 |
| **Time since last COVID monovalent vaccination** |  |  | 0.2013 |  |  | 0.0781 |
| <= 90 days | 540 (0.4%) | 3,820 (3.0%) |  | 1,065 (0.9%) | 2,182 (1.7%) |  |
| 91-180 days | 11,305 (9.0%) | 11,478 (9.1%) |  | 11,372 (9.1%) | 11,393 (9.0%) |  |
| 180+ days | 95,595 (75.8%) | 92,107 (73.0%) |  | 93,622 (75.0%) | 93,693 (74.4%) |  |
| Not reported | 18,696 (14.8%) | 18,731 (14.8%) |  | 18,803 (15.1%) | 18,699 (14.8%) |  |
| **Time since last COVID infection** |  |  | 0.0317 |  |  | 0.0018 |
| <= 120 days | 269 (0.2%) | 282 (0.2%) |  | 273 (0.2%) | 275 (0.2%) |  |
| 121-180 days | 447 (0.4%) | 418 (0.3%) |  | 430 (0.3%) | 430 (0.3%) |  |
| 180+ days | 3,884 (3.1%) | 3,231 (2.6%) |  | 3,512 (2.8%) | 3,497 (2.8%) |  |
| Not reported | 121,536 (96.4%) | 122,205 (96.9%) |  | 120,647 (96.6%) | 121,764 (96.7%) |  |
| **Evidence of COVID vaccination with prior season's vaccine during the pre-index period** | 107,367 (85.1%) | 107,367 (85.1%) |  | 106,001 (84.9%) | 107,219 (85.1%) | 0.0062 |
| **Evidence of influenza vaccination during the pre-index period** | 114,756 (91.0%) | 105,687 (83.8%) | 0.2178 | 109,026 (87.3%) | 109,951 (87.3%) | 0.0009 |
| **Diagnosis of influenza during pre-index period** | 2,331 (1.8%) | 2,561 (2.0%) | 0.0132 | 2,431 (1.9%) | 2,450 (1.9%) | 0.0002 |
| **Week of last healthcare activity before start of intake** |  |  |  |  |  | 0.0077 |
| 0-4 weeks | 117,438 (93.1%) | 117,438 (93.1%) |  | 116,268 (93.1%) | 117,321 (93.1%) |  |
| 5-26 weeks | 8,596 (6.8%) | 8,596 (6.8%) |  | 8,490 (6.8%) | 8,542 (6.8%) |  |
| 27-52 weeks | 69 (0.1%) | 69 (0.1%) |  | 69 (0.1%) | 69 (0.1%) |  |
| > 52 weeks or unknown | 33 (0.0%) | 33 (0.0%) |  | 35 (0.0%) | 33 (0.0%) |  |

SMD = standardized mean difference. Values with absolute SMD >0.10 indicate covariate imbalance. Post-weighting values (IPTW-adjusted) are used in Table 1 characteristics; pre-weighting patient counts are used in all text. Vaccine counts (n) in the comorbidity rows reflect presence of the condition (=1).

**Supplementary Table 7.** Baseline Characteristics (Pre-weighting and Post-weighting, and SMDs) – BNT162b2 ≥75 Years.

Pre-weighting N: 227,989 vaccinated / 227,989 comparator | Post-weighting N: 225,863 vaccinated / 227,746 comparator
|  | Pre-weighting |  |  | Post-weighting (IPTW-adjusted) |  |  |
| --- | --- | --- | --- | --- | --- | --- |
|  | Vaccinated | Comparator | SMD | Vaccinated | Comparator | SMD |
| **Number of Patients** | 227,989 | 227,989 |  | 225,863 | 227,746 |  |
| **Age; Mean (SD)** | 81 (4.3) | 81 (4.3) |  | 81 (4.2) | 81 (4.3) | 0.0036 |
| **Age Categories; n (%)** |  |  |  |  |  | 0.0018 |
| 75-79 | 105,977 (46.5%) | 105,977 (46.5%) |  | 104,687 (46.3%) | 105,719 (46.4%) |  |
| 80-84 | 69,025 (30.3%) | 69,025 (30.3%) |  | 68,398 (30.3%) | 68,956 (30.3%) |  |
| 85-89 | 52,987 (23.2%) | 52,987 (23.2%) |  | 52,778 (23.4%) | 53,071 (23.3%) |  |
| **Sex; n (%)** |  |  |  |  |  | 0.0003 |
| Male | 98,873 (43.4%) | 98,873 (43.4%) |  | 97,951 (43.4%) | 98,802 (43.4%) |  |
| **Race; n (%)** |  |  |  |  |  | 0.0009 |
| Black | 15,423 (6.8%) | 15,423 (6.8%) |  | 15,291 (6.8%) | 15,372 (6.7%) |  |
| White | 148,971 (65.3%) | 148,971 (65.3%) |  | 147,612 (65.4%) | 148,914 (65.4%) |  |
| Other | 31,307 (13.7%) | 31,307 (13.7%) |  | 31,048 (13.7%) | 31,285 (13.7%) |  |
| Unknown | 32,288 (14.2%) | 32,288 (14.2%) |  | 31,911 (14.1%) | 32,176 (14.1%) |  |
| **Ethnicity; n (%)** |  |  |  |  |  | 0.0007 |
| Hispanic | 4,385 (1.9%) | 4,385 (1.9%) |  | 4,371 (1.9%) | 4,385 (1.9%) |  |
| Non-Hispanic | 136,237 (59.8%) | 136,237 (59.8%) |  | 134,989 (59.8%) | 136,114 (59.8%) |  |
| Unknown | 87,367 (38.3%) | 87,367 (38.3%) |  | 86,504 (38.3%) | 87,248 (38.3%) |  |
| **Insurance Type; n (%)** |  |  | 0.0999 |  |  | 0.0055 |
| Commercial | 3,330 (1.5%) | 3,110 (1.4%) |  | 3,195 (1.4%) | 3,219 (1.4%) |  |
| Medicaid | 879 (0.4%) | 1,983 (0.9%) |  | 1,332 (0.6%) | 1,435 (0.6%) |  |
| Medicare Advantage | 200,146 (87.8%) | 193,998 (85.1%) |  | 195,398 (86.5%) | 196,816 (86.4%) |  |
| Medicare FFS | 18,837 (8.3%) | 22,102 (9.7%) |  | 20,265 (9.0%) | 20,500 (9.0%) |  |
| Unknown | 4,797 (2.1%) | 6,796 (3.0%) |  | 5,672 (2.5%) | 5,776 (2.5%) |  |
| **Region; n (%)** |  |  |  |  |  | 0.0015 |
| Midwest | 40,319 (17.7%) | 40,319 (17.7%) |  | 40,077 (17.7%) | 40,333 (17.7%) |  |
| Northeast | 49,367 (21.7%) | 49,367 (21.7%) |  | 48,973 (21.7%) | 49,354 (21.7%) |  |
| South | 83,851 (36.8%) | 83,851 (36.8%) |  | 83,017 (36.8%) | 83,664 (36.7%) |  |
| West | 54,448 (23.9%) | 54,448 (23.9%) |  | 53,792 (23.8%) | 54,390 (23.9%) |  |
| Unknown | 4 (0.0%) | 4 (0.0%) |  | 4 (0.0%) | 4 (0.0%) |  |
| **Month of Index Date; n (%)** |  |  |  |  |  | 0.0067 |
| 2025-08 | 86 (0.0%) | 86 (0.0%) |  | 89 (0.0%) | 84 (0.0%) |  |
| 2025-09 | 87,613 (38.4%) | 87,613 (38.4%) |  | 86,795 (38.4%) | 87,129 (38.3%) |  |
| 2025-10 | 96,659 (42.4%) | 96,659 (42.4%) |  | 95,766 (42.4%) | 96,428 (42.3%) |  |
| 2025-11 | 32,106 (14.1%) | 32,106 (14.1%) |  | 31,772 (14.1%) | 32,365 (14.2%) |  |
| 2025-12 | 10,176 (4.5%) | 10,176 (4.5%) |  | 10,124 (4.5%) | 10,332 (4.5%) |  |
| 2026-01 | 1,349 (0.6%) | 1,349 (0.6%) |  | 1,317 (0.6%) | 1,408 (0.6%) |  |
| **Number of OP Visits; Mean (SD)** | 2.6 (6.1) | 2.8 (6.6) | 0.0290 | 2.7 (6.3) | 2.7 (6.3) | 0.0011 |
| **Number of Hospitalizations; Mean (SD)** | 0.1 (0.5) | 0.2 (0.5) | 0.0808 | 0.2 (0.5) | 0.2 (0.5) | 0.0020 |
| **Baseline Clinical Comorbidities; n (%)** |  |  |  |  |  |  |
| Cancer | 41,572 (18.2%) | 40,407 (17.7%) | 0.0133 | 40,633 (18.0%) | 40,955 (18.0%) | 0.0002 |
| Cerebrovascular disease | 36,055 (15.8%) | 41,398 (18.2%) | 0.0624 | 38,292 (17.0%) | 38,720 (17.0%) | 0.0013 |
| Chronic kidney disease | 77,216 (33.9%) | 81,539 (35.8%) | 0.0398 | 78,713 (34.9%) | 79,377 (34.9%) | 0.0001 |
| Chronic lung diseases | 46,442 (20.4%) | 52,346 (23.0%) | 0.0629 | 48,942 (21.7%) | 49,411 (21.7%) | 0.0006 |
| Chronic liver diseases | 3,469 (1.5%) | 4,059 (1.8%) | 0.0203 | 3,707 (1.6%) | 3,756 (1.6%) | 0.0006 |
| Cystic fibrosis | 29 (0.0%) | 24 (0.0%) | 0.0020 | 26 (0.0%) | 25 (0.0%) | 0.0004 |
| Diabetes mellitus, type 1 and type 2 | 75,682 (33.2%) | 84,304 (37.0%) | 0.0793 | 79,354 (35.1%) | 80,066 (35.2%) | 0.0005 |
| Disabilities | 11,971 (5.3%) | 11,665 (5.1%) | 0.0061 | 11,696 (5.2%) | 11,794 (5.2%) |  |
| Heart conditions | 88,882 (39.0%) | 95,726 (42.0%) | 0.0612 | 91,437 (40.5%) | 92,305 (40.5%) | 0.0009 |
| HIV | 375 (0.2%) | 308 (0.1%) | 0.0076 | 339 (0.2%) | 339 (0.1%) | 0.0003 |
| Mental health disorders | 50,009 (21.9%) | 57,047 (25.0%) | 0.0729 | 52,981 (23.5%) | 53,524 (23.5%) | 0.0010 |
| Neurologic conditions | 25,110 (11.0%) | 34,434 (15.1%) | 0.1216 | 29,474 (13.0%) | 29,812 (13.1%) | 0.0012 |
| Obesity | 54,789 (24.0%) | 57,959 (25.4%) | 0.0322 | 55,901 (24.7%) | 56,400 (24.8%) | 0.0003 |
| Primary Immunodeficiencies | 6,770 (3.0%) | 6,969 (3.1%) | 0.0051 | 6,790 (3.0%) | 6,851 (3.0%) | 0.0001 |
| Physical inactivity | 438 (0.2%) | 553 (0.2%) | 0.0108 | 488 (0.2%) | 496 (0.2%) | 0.0004 |
| Smoking, current and former | 49,501 (21.7%) | 53,119 (23.3%) | 0.0380 | 50,776 (22.5%) | 51,276 (22.5%) | 0.0008 |
| Solid organ or hematopoietic cell transplantation | 518 (0.2%) | 541 (0.2%) | 0.0021 | 519 (0.2%) | 525 (0.2%) | 0.0002 |
| Tuberculosis | 95 (0.0%) | 146 (0.1%) | 0.0097 | 120 (0.1%) | 122 (0.1%) | 0.0002 |
| Use of immunosuppressive medications | 18,066 (7.9%) | 15,885 (7.0%) | 0.0364 | 16,835 (7.5%) | 16,938 (7.4%) | 0.0006 |
| Asthma | 20,696 (9.1%) | 20,002 (8.8%) | 0.0107 | 20,195 (8.9%) | 20,339 (8.9%) | 0.0004 |
| **Time since last COVID monovalent vaccination** |  |  | 0.1911 |  |  | 0.0773 |
| <= 90 days | 892 (0.4%) | 6,177 (2.7%) |  | 1,646 (0.7%) | 3,538 (1.6%) |  |
| 91-180 days | 15,683 (6.9%) | 16,847 (7.4%) |  | 16,116 (7.1%) | 16,204 (7.1%) |  |
| 180+ days | 174,461 (76.5%) | 167,948 (73.7%) |  | 170,960 (75.7%) | 171,037 (75.1%) |  |
| Not reported | 36,953 (16.2%) | 37,017 (16.2%) |  | 37,141 (16.4%) | 36,968 (16.2%) |  |
| **Time since last COVID infection** |  |  | 0.0221 |  |  | 0.0012 |
| <= 120 days | 496 (0.2%) | 574 (0.3%) |  | 520 (0.2%) | 530 (0.2%) |  |
| 121-180 days | 770 (0.3%) | 727 (0.3%) |  | 738 (0.3%) | 744 (0.3%) |  |
| 180+ days | 6,561 (2.9%) | 5,791 (2.5%) |  | 6,100 (2.7%) | 6,104 (2.7%) |  |
| Not reported | 220,162 (96.6%) | 220,897 (96.9%) |  | 218,506 (96.7%) | 220,368 (96.8%) |  |
| **Evidence of COVID vaccination with prior season's vaccine during the pre-index period** | 190,889 (83.7%) | 190,889 (83.7%) |  | 188,605 (83.5%) | 190,676 (83.7%) | 0.0059 |
| **Evidence of influenza vaccination during the pre-index period** | 206,675 (90.7%) | 189,010 (82.9%) | 0.2302 | 195,865 (86.7%) | 197,445 (86.7%) | 0.0007 |
| **Diagnosis of influenza during pre-index period** | 4,493 (2.0%) | 4,757 (2.1%) | 0.0082 | 4,593 (2.0%) | 4,626 (2.0%) | 0.0002 |
| **Week of last healthcare activity before start of intake** |  |  |  |  |  | 0.0083 |
| 0-4 weeks | 212,730 (93.3%) | 212,730 (93.3%) |  | 210,756 (93.3%) | 212,591 (93.3%) |  |
| 5-26 weeks | 15,078 (6.6%) | 15,078 (6.6%) |  | 14,928 (6.6%) | 14,978 (6.6%) |  |
| 27-52 weeks | 132 (0.1%) | 132 (0.1%) |  | 130 (0.1%) | 129 (0.1%) |  |
| > 52 weeks or unknown | 49 (0.0%) | 49 (0.0%) |  | 49 (0.0%) | 48 (0.0%) |  |

SMD = standardized mean difference. Values with absolute SMD >0.10 indicate covariate imbalance. Post-weighting values (IPTW-adjusted) are used in Table 1 characteristics; pre-weighting patient counts are used in all text. Vaccine counts (n) in the comorbidity rows reflect presence of the condition (=1).

